# Risk factors for food contamination among children 6-59 months discharged from community management of acute malnutrition (CMAM) programmes for severe acute malnutrition (SAM) in Aweil East, South Sudan

**DOI:** 10.1101/2023.04.14.23288551

**Authors:** Joseph Wells, David Gama Abugo, John Angong, Nancy Grace Lamwaka, Karin Gallandat, Jackson Lwate Hassan, Lino Deng, Dimple Save, Laura Braun, Mesfin Gose, Jacob Amanya, Khamisa Ayoub, Sarah King, Heather Stobaugh, Oliver Cumming, Lauren D’Mello-Guyett

## Abstract

Children under-five years of age are particularly vulnerable to severe acute malnutrition (SAM), and the risk factors associated with relapse to SAM are poorly understood. Possible causes are asymptomatic or symptomatic infection with enteric pathogens, with contaminated food as a critical transmission route. This cross-sectional study comprised a household survey with samples of child food (n=382) and structured observations of food preparation (n=197) among children aged 6-59 months that were discharged from treatment in community management of acute malnutrition (CMAM) programmes in Aweil East, South Sudan. We quantified *Escherichia coli* and total faecal coliforms (TFCs), measured in colony forming units per g of food (CFU/g), as indicators of microbial contamination of child food. A modified hazard analysis critical control point (HACCP) approach was utilised to determine critical control points (CCPs) followed by multivariate logistic regression analysis to understand the risk factors associated with contamination. Over 40% of samples were contaminated with *E. coli* (43% >0 *E. coli* CFU/g, 95%CI 38-48%), and 90% had >10 TFCs (CFU/g) (>10TFC CFU/g, 95%CI 87-93%). Risk factors associated (p<0.05) with child food contamination included if the child fed themselves (95% CI 1.16, 3.44, log[odds] = 2.20), exposure to animals (95% CI 0.28, 1.68, log[odds] = 0.97), and protective factors for contamination included feeding with a spoon (95% CI -7.81, -1.54, log[odds] = -4.17). This study highlights strategies that can support interventions that reduce food contamination exposure in young children and help further protect those that are highly vulnerable to recurrent exposure to diarrhoeagenic pathogens.

## Introduction

In 2020, estimates suggest 13.6 million children under the age of five suffered from severe acute malnutrition (SAM) (WHO, 2021). Over 300,000 of these estimated cases were from children under-five in South Sudan (UNICEF, 2022). SAM is a short-term acute condition with a high case-fatality rate (CFR) that increases vulnerability to infections and disease (Olofin et al., 2013). There is a significant body of research addressing the risk factors, short-term health implications and treatment for SAM (O’Sullivan, Lelijveld, Rutishauser-Perera, Kerac, & James, 2018). However, little is known about children’s health and nutrition following discharge from SAM/ moderate acute malnutrition (MAM) treatment programmes or why some children are more at risk of relapse than others (Stobaugh et al., 2019).

Globally, relapse to SAM within the first six months post-treatment ranges from 0% to 37%, whilst the mortality rate ranges from 0.1% to 10.4% within 12 months post-discharge (Stobaugh et al., 2019). Evidence has suggested that severe and persistent diarrhoeal disease among children post-SAM is a key factor for relapse (Adegoke et al., 2021). In Bangladesh, (Ashraf et al., 2012) 20% of children reported diarrhoea in the two-week post-discharge period, which decreased to 6% at three months post-discharge. Gastrointestinal disease is also likely to be underreported as enteric infections are often asymptomatic (Cichon et al., 2016). This highlights that there may be an association between enteric infections, whether symptomatic or asymptomatic and an increased risk of relapse.

Food is an important route of transmission for enteric diseases in children under-five years of age (Havelaar et al., 2015). These infections can be symptomatic or asymptomatic but commonly present as gastrointestinal syndromes – with symptoms such as diarrhoea, nausea and vomiting (Havelaar et al., 2015). Enteric pathogens can weaken a child’s immune system; impact their cognitive, motor and social-emotional development (Kotloff et al., 2013; Ngure et al., 2013); and, lead to subsequent or exacerbated malnutrition (Petri et al., 2008; Schaible & Kaufmann, 2007). Crucially, these infections are likely to be a risk factor for relapse in children who have recovered from SAM.

Globally, there are an estimated 600 million cases of foodborne illnesses each year, with children under-five bearing 40% of this burden (Havelaar et al., 2015). In East Africa, *Salmonella*, cholera and toxigenic *E. coli* are the three diarrheagenic foodborne diseases that are responsible for the greatest loss of disability-adjusted life years (DALYs) (WHO, 2015). Multiple studies in low- and middle-income countries (LMICs) have found high levels of microbial contamination in child foods (Bick et al., 2020; Ercumen et al., 2017; Gizaw, Yalew, Bitew, Lee, & Bisesi, 2022; Islam et al., 2013; Touré, Coulibaly, Arby, Maiga, & Cairncross, 2011; Tsai et al., 2019). For example, *E. coli* was detected in 68% of child food samples in rural Ethiopia (Gizaw et al., 2022) and 48% of samples in rural Malawi (Taulo et al., 2008).

Modified hazard analysis and critical control point (HACCP) methods are recognised as an effective tool for understanding and preventing food contamination in LMIC households (Bick et al., 2020; Gautam & Curtis, 2021; Islam et al., 2013; Manjang et al., 2018; Touré et al., 2011). HACCP is a management system that addresses food safety through the analysis of food product information and assessment of physical, chemical, and biological hazards throughout food production processes. Critical control points (CCPs) are assigned to steps in the process at which controls could be applied to reduce, prevent or eliminate hazards to an acceptable level (FAO & WHO Codex Alimentarius Commission, 1997).

Studies employing HACCP and other quantitative approaches to explore child food contamination and hygiene in LMICs have identified common CCPs. Poor handwashing with soap (HWWS) practices (Bick et al., 2020; Davis et al., 2018; Islam et al.,2013), household animal ownership and the presence of animal waste (Barnes, Anderson, Mumma, Mahmud, & Cumming, 2018; Ercumen et al., 2017; Parvez et al., 2017) and poor storage practices (Ercumen et al., 2017; Touré et al., 2011) have all been identified as risk factors for child food contamination in both urban and rural contexts. However, the risk of contamination in food fed to children that have suffered and recovered from SAM in humanitarian settings has yet to be studied.

To design and implement effective water, sanitation and hygiene (WASH) strategies to improve food hygiene, it is critical to understand the complex pathways through which food may become contaminated within households. This exploratory, cross-sectional study aimed to assess the prevalence and quantity of *E. coli* and total faecal coliform (TFC) contamination, as indicators for faecal contamination, in child foods for children that are at risk of relapse to SAM. Additionally, the study aimed to identify associated risk factors for food contamination. We applied a modified HACCP approach, paired with a risk factor analysis, to explore potential practices and risk factors that would contribute to food contamination in a low-income setting of Aweil East, South Sudan. The findings of this study can support the design and implementation of strategies to reduce exposure to foodborne contamination.

## Key messages

- Food is a potentially important pathway for enteric disease transmission and a major concern for children recovered from severe acute malnutrition (SAM) in humanitarian settings.
- In low-income Aweil East, South Sudan, 43% of child food samples from households with a child at risk of relapse to SAM were contaminated with *E. coli* and 90% with total faecal coliforms (TFCs).
- Child self-feeding and animal presence in the household were identified as key risk factors for food contamination.
- Future research should assess the relative importance of the identified critical control points (CCPs), including reducing animal contact in the household, improving knowledge and practices around cooking processes and hygiene practices specific to the child such as handwashing and the use of cutlery.

## Methods

### Study Design, Setting and Population

This was an exploratory, cross-sectional study, nested within a multi-country prospective cohort study of children following anthropometric recovery from SAM aged 6-59 months in South Sudan, Somalia and Mali (King et al., 2022). The study was a collaboration between the London School of Hygiene and Tropical Medicine (LSHTM), Action Against Hunger (ACF) and the Ministry of Health in South Sudan.

Aweil East, South Sudan, is in Northern Bahr el Ghazal State which contains seven payams, with a total population of 538,765 of which 102,365 are under-five years of age. A 2021 SMART survey identified a Global Acute Malnutrition (GAM) and SAM prevalence of 13.1% and 2.6%, respectively (ACF, 2021). The county is routinely classified as IPC Phase 3 (Crisis) or IPC Phase 4 (Emergency), signifying widespread food insecurity and high rates of acute malnutrition. The region was classified as Phase 4 in 2022 (IPC, 2022). Aweil East has a total of 13 government health facilities and outreach clinics in the area, all of which are supported by ACF. Each health facility is approximately 20-30 km apart. Six out of the 13 health facilities were included in the study, with sites in urban, peri-urban and rural settings.

The study enrolled children aged 6-59 months who have recovered from SAM at the point of discharge from treatment within a CMAM programme (discharge was defined as recovered according to the global WHO definition of recovery from SAM (WHO, 2013): with a mid-upper arm circumference (MUAC) ≥ 125 mm, weight-for-height Z-score (WHZ) ≥ -2 and/or no oedema for two consecutive weeks).

### Sample size calculation

The study enrolled 618 children discharged from CMAM programmes in Aweil East, South Sudan. Using a power of 0.8, a level of significance of 0.05, and assuming the true proportion relapsing within 12 months of discharge to be 12% (Stobaugh et al., 2019), the minimum detectable differences (MDD) between exposed and unexposed groups (e.g., proportion with/without adequate household WASH, proportion with/without enteric pathogens detected, and proportion with/without contaminated food and drinking water) under different exposure scenarios (i.e., proportion exposed from 10% – 80%) were calculated using the R package “powerMediation”, function “SSizeLogisticBin”. A sample size of 618 would be adequate to detect statistically significant differences in the proportion relapsing of approximately at least 20 percentage points between groups and under different exposure scenarios. For the selection of households to include in the structured observations, we selected every third child enrolled on the study.

### Data collection: household survey and structured observations

We conducted a household survey in 618 households to characterise sociodemographic factors, asset ownership, WASH conditions (including animal contact and exposure, hygiene and handwashing, sanitation, child food faeces disposal, water access, water treatment and storage), child intake, and food preparation and storage practices (Appendix A: Figure A1). Trained data collectors administered the survey via a home visit within two weeks of discharge from CMAM programmes and as soon as possible after enrolment into the study.

In 197 of the 618 households, we conducted additional structured observations of food preparation, storage and hygiene practices (Appendix A: Figure A1). Trained enumerators conducted observations of caregivers to record types of food served, cleaning and washing of ingredients, utensils, preparation/feeding surfaces and cups, food additions before and after cooking, reheating practices, separation of cooked and raw foods, HWWS of caregivers and children, how the child is fed including child seating position, food storage areas, and, animal contact during food preparation and feeding. Data collectors arranged a time with the caregiver to observe the preparation and serving of the child’s food. Observations lasted between 1-3 hours, depending on the duration of preparation, feeding and food storage activities.

The survey and structured observations were recorded on tablets using Open Data Kit (ODK) submitted to and checked by the Field Research Coordinator, who uploaded the data to secure ODK servers at LSHTM. All data collection was conducted between June 2021 and October 2022.

### Data collection: food sample collection and analysis

Alongside the household survey and structured observations, enumerators asked caregivers to provide a small amount of food they would give to the child. Samples were collected according to standardised and pre-tested protocols. An estimated 10g of the child’s food was placed in sterile 100ml WhirlPak® bags and transported on ice to a field laboratory to be processed within 6 hours of collection. If meals had multiple food components, samples of all components were homogenised by shaking the bags. Following standard Association of Official Analytical Chemists (AOAC) methods (#991.14) (Da Silva et al., 2018), a known amount of each food sample (11 g) was blended with 89 g of saline solution (0.9% Sodium chloride (NaCl)). The pH was adjusted to 6.6-7.2, with either hydrochloric acid (HCl) or sodium hydroxide (NaOH), and 1 mL of the diluted sample was placed onto a 3M^TM^ PetriFilm^TM^ for the simultaneous detection of the primary outcome, *E. coli* and secondary outcome, total faecal coliform*s* (TFCs), as indicators of faecal contamination (Paruch & Mæhlum, 2012). Bacteria were enumerated as colony-forming units (CFU) per gram (CFU/g) of the food sample. The limit of detection for 3MTM PetriFilm^TM^ is 1 CFU/g. Negative controls and duplicates were analysed in each batch of samples.

### Modified HACCP analysis methodology

HACCP is a management system in which food safety is addressed through analysing food product information and assessing physical, chemical and biological hazards throughout the product’s lifetime. Critical limits are specified, and CCPs are identified (FAO & WHO Codex Alimentarius Commission, 1997) along the food preparation process.

A modified HACCP analysis was conducted on the observation data. All foods were categorised based on ingredients and preparation processes (FAO & WHO, 2017). A food-flow diagram was constructed for each food category based on the information collected from the structured observation data. A general food-flow diagram was also constructed for all foods. Descriptive statistics were generated to describe common preparation, feeding and storage practices but also compare these practices between food groups. Evaluation of the food preparation process was used to identify a range of points that may impact the prevalence or magnitude of contamination. Control measures, if any, were then identified for each hazard. CCPs were then identified out of the range of control points using a decision tree and based on whether (a) preventive control measures can be applied, (b) the step specifically eliminates or reduces the occurrence of hazard to an acceptable level, (c) loss of control could increase contamination to unacceptable levels, and (d) subsequent step eliminates identified hazard(s) or reduces occurrence to an acceptable level (FAO & WHO Codex Alimentarius Commission, 1997). Food-flow diagrams were annotated with CCPs, and feasible control measures were suggested to reduce *E. coli* and TFC contamination in child foods.

### Statistical analysis

All statistical analyses were conducted in RStudio 2022.02.2 Build 485 “Prairie Trillium” (R Foundation for Statistical Computing, Vienna, Austria).

To estimate relative wealth, sociodemographic, asset and wealth variables, commonly used in the Demographic Health Survey (DHS), were used to create a wealth index based on principal component analysis (*FactoMineR* package*: ‘PCA’* function). 31 variables were used to construct the index: asset ownership (12 binary variables), household size (continuous), number of rooms (continuous), housing characteristics (8 binary variables), animal ownership (binary), and ownership of different animal species (8 continuous variables).

Housing characteristics including roof, wall and floor materials were dichotomised into low- and high-quality construction materials (Khudri & Chowdhury, 2013). High-quality construction materials included concrete, laminated wood or tiles. Low-quality construction materials were associated with poorer structural stability and less easy to clean, such as mud, thatch or wooden planks. The exclusion of households based on missing socio-economic data can significantly impact sample size and statistical power, so missing values were statistically imputed (*missMDA* package*: ‘MIFAMD’* function) (Vyas & Kumaranayake, 2006). The first principal component analysis was used to group the households by socioeconomic status (SES) into relative wealth quintiles.

Arithmetic means were used to report colony counts (Haas, 1996). For *E. coli*, an outcome of >0 CFU/g was defined as “contaminated”, while 0 CFU/g was defined as “non-contaminated”. Guideline values for acceptable *E. coli* in food vary depending on the product and we decided conservatively to classify no detected *E. coli* as safe (Hasell & Salter, 2003). For TFC, an outcome of >10 CFU/g was defined as “contaminated”, while ≤10 CFU/g was defined as “non-contaminated” as other studies have reported (Touré, Coulibaly, Arby, Maiga, & Cairncross, 2013). The prevalence of *E. coli* and TFC contamination in child foods was calculated alongside 95% confidence intervals (CIs). The proportion of samples contaminated was calculated with logic transformed 95% confidence intervals.

All other variables were converted to binary or categorical variables, and food types were grouped based on common ingredients of samples and food preparation processes. Households were categorised into the WHO/UNICEF Joint Monitoring Programme (JMP) ladder definitions for drinking water, sanitation and hygiene (WHO & UNICEF, 2023).

Univariate and multivariable analysis was conducted to test for associations between variables and the primary and secondary outcomes. Individual variables were tested for association with contamination using univariate binomial logistical regression (*stats* package*: ‘glm’* function). Variables with a *p* value below 0.1 in the binomial logistical regression and those variables that perfectly predicted the outcome were considered for inclusion in the binomial portion of the multivariable model. For each iteration of the multivariable model, variables were added individually in a stepwise procedure. Akaike Information Criterion (AIC) was used to compare the base model (Null model) with each new model. The variable was retained where the statistic was minimised by 1 AIC (ΔAIC = 1) but if the variable’s *p* value scored >0.1, it was not retained. A likelihood ratio test (*lmtest* package: ‘lrtest’ function) was also used to understand whether each model was significantly improved, compared with the previous model. During each step, variables were checked to ensure they continued to have a *p* value < 0.1, or otherwise, they were removed from the next step. The process was repeated until no variables improved the model fit and all scored a *p* value *< 0.1*. Age, gender and SES were added to the model as confounders. The final multivariable model was checked for multicollinearity, with variables exceeding a variance inflation factor of 10 removed (*car* package*: ‘vif’* function). Variables with a final *p* value *< 0.05* were classed as significant.

### Ethical considerations

Ethical clearance was obtained both before the enrolment of study participants and before the analysis of child food contamination data from LSHTM (LSHTM MSc Ethics Ref: 18059), Ministry of Health in South Sudan (No. 02/06/2020-MoH/RERB/A/01/2020) and Solutions IRB (No. #2020/03/10). Informed written consent was obtained from all study participants.

## Results

### Characteristics of study participants and exposures

A total of 618 households were included in the study, however, based on the availability of food in the households, we were only able to collect food samples from 382 of the 618 households enrolled (Appendix A: Figure A1).

Of these 382 households, caregivers responsible for preparing and feeding the child were predominantly female (94%). The children in the study were 52% male and 48% female, with a mean age of 34 months. The median household size was 7 (IQ range: 6-9), and the average number of rooms in the house was 2 (IQ range: 1-3). The floor (99%), walls (98%) and roof (98%) of houses were predominantly made of low-quality materials such as mud, thatch or wood planks. Households were split into the five wealth quintiles (1^st^ = highest wealth quintile, 5^th^ = lowest wealth quintile), with the highest proportion in the 1^st^ quintile (23%) and the lowest proportion in the 5^th^ quintile (16%).

### Water, sanitation and hygiene

The majority of households had access to a basic drinking water service (79%), meaning that they were using an improved source with a roundtrip collection time of no more than 30 minutes including queueing. Few (24%) respondents reported water was treated before consumption, with cloth filtration the most popular treatment practice (45%). 39% of household drinking water was stored with a cover or lid. All stored drinking water samples had a median free residual chlorine concentration of <0.5mg/L, indicating that water is likely not appropriately treated with chlorine, while 38% had a turbidity of more than 5 nephelometric turbidity units (NTU). Of the 379 drinking water samples, 80% had >0 *E. coli* (CFU/100ml) and 98% were contaminated with >10 TFC (CFU/100ml). On average, households had 55 litres (SD = 40) of water in the house of which 33 litres (SD = 26) were reserved for drinking water. The majority (59%) of households had not experienced insufficient drinking water in the household within the past four weeks, but 32% had experienced water scarcity once or twice in this period. The main reason for experiencing water scarcity was that the water point needed to be fixed (77%). The majority of households (60%) had an alternative source of drinking water.

For sanitation, the majority (77%) of households reported open defecation (disposal of faeces in the environment e.g., fields, forests), with 14% reporting the use of unimproved facilities (pit latrine without a platform, hanging latrine or bucket latrine) and 9% limited facilities (improved facility, shared between two or more households). Child faeces were frequently disposed of outside the house in the open (80%), and in some cases, there were human faeces observed inside the compound (14%).

The majority (75%) had limited handwashing facilities (available handwashing facility lacking soap and/or water), 20% of households had a basic facility (available handwashing facility with soap and water present), and 5% had no facility whatsoever. Soap was present in 47% of households, with less than half (45%) of survey respondents using soap and water to wash their hands. Only 28% of households reported HWWS at the five key times (after defecation, after changing a child’s nappy/diaper/faeces, before food preparation, before eating and after eating). Garbage in the compound was also observed in 69% of households.

### Animal exposures

Households reported they were most commonly exposed to chickens (54%), goats/sheep (29%) and cattle (17%). In 36% of households, animal faeces were observed in the compound. During food preparation and feeding, chickens (54%), goats/sheep (15%) and dogs (10%) were most commonly observed in food preparation or feeding areas.

### Food preparation

The majority of households washed their fruit and vegetables before cooking or feeding their child. The most common time to cook food was in the late evening, after sunset (45%). Only 59% of households covered stored food. A minority of households (34%) reported eating leftovers, leftovers were commonly not re-cooked (49%) whereas some households reported boiling their leftovers (32%).

Of the 197 structured observations, the three most common ingredients of food fed to children included cereals (98%), milk (27%) and fish (17%). Water (83%) was added most frequently before or during cooking. When something was added to food, the additional ingredient was heated only 39% of the time. Food was most commonly prepared directly in a cooking pot (60%) compared to on the table (2%) or floor (12%).

All exposures are reported in Tables 1 and 2.

**Table 1.**
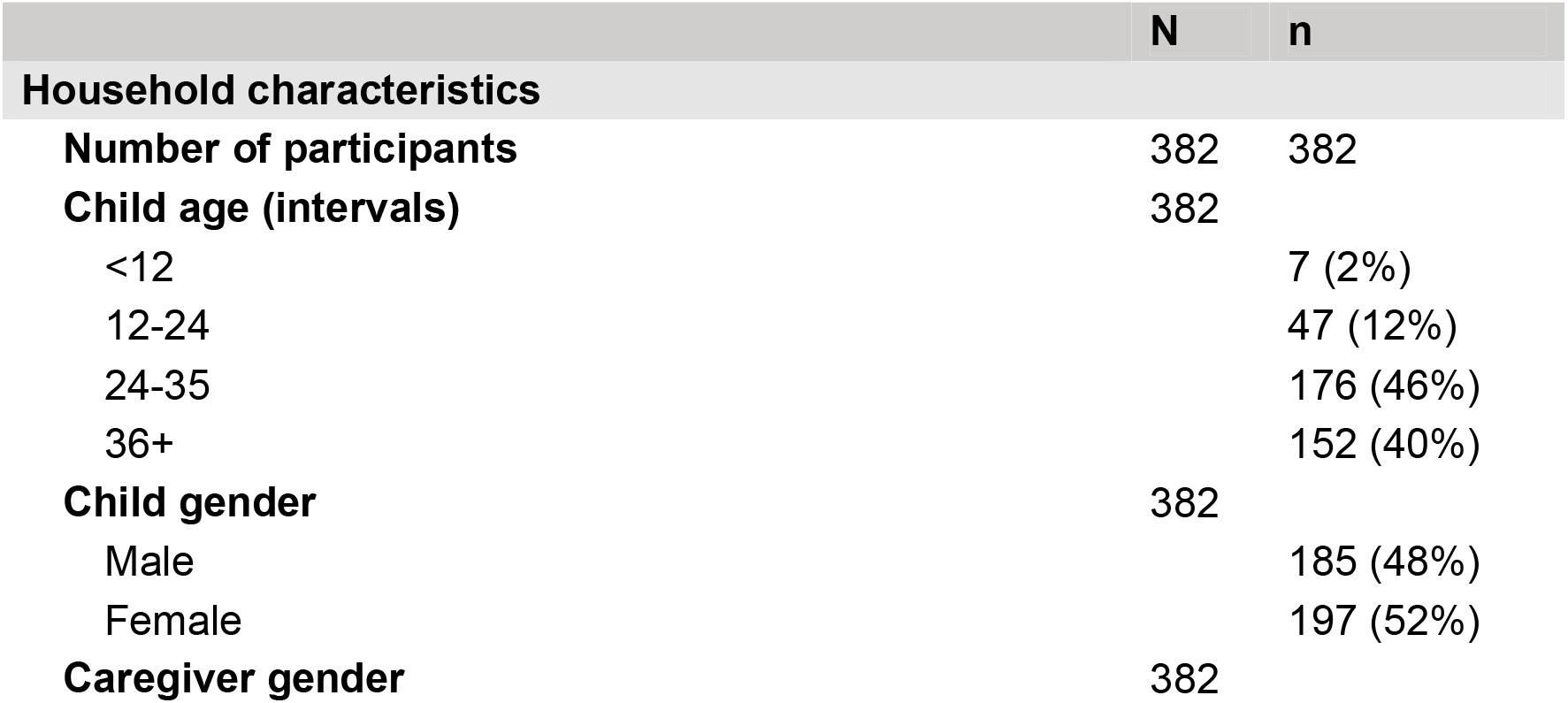
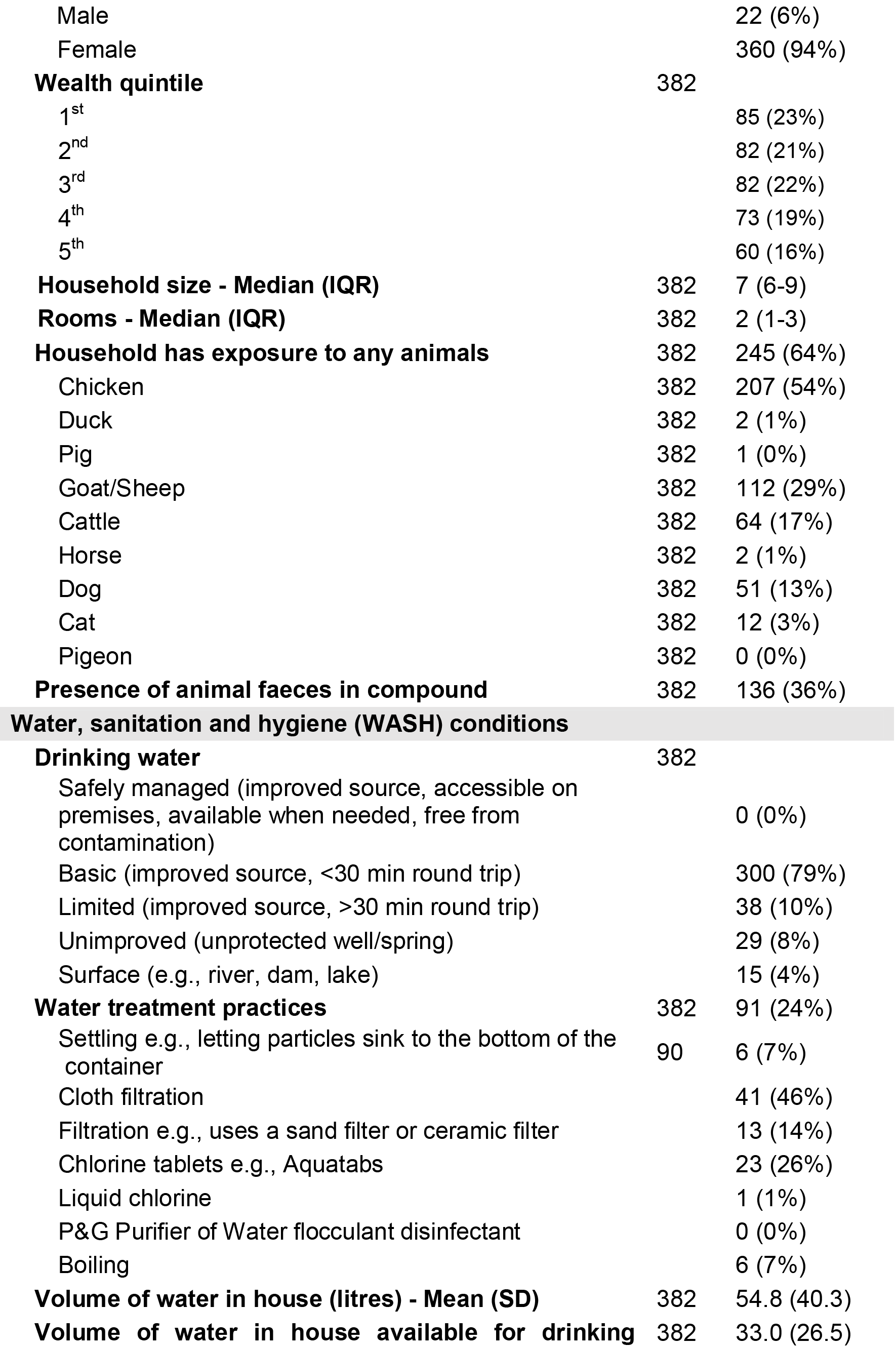
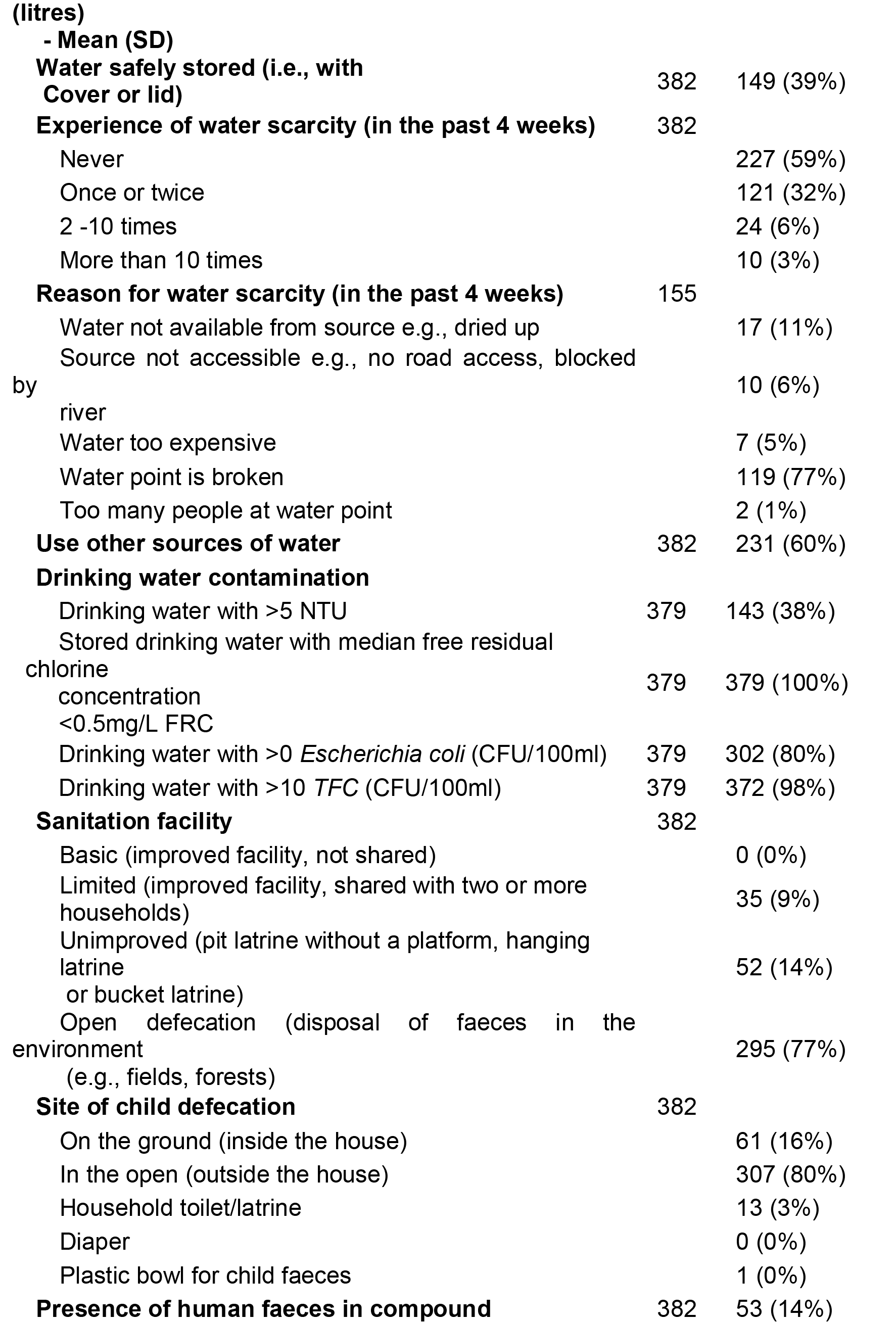
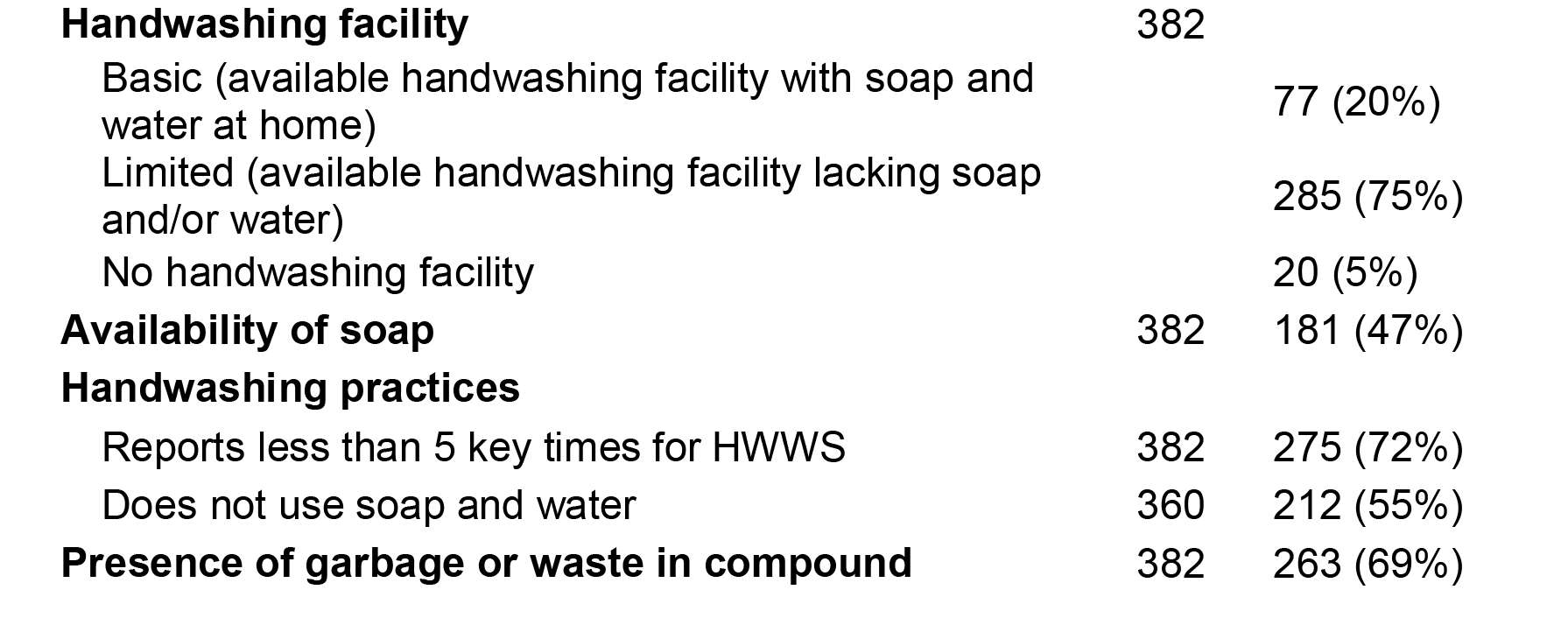
Characteristics of 382 households of children recovered from severe acute malnutrition (SAM), Aweil East, South Sudan.

**Table 2.**
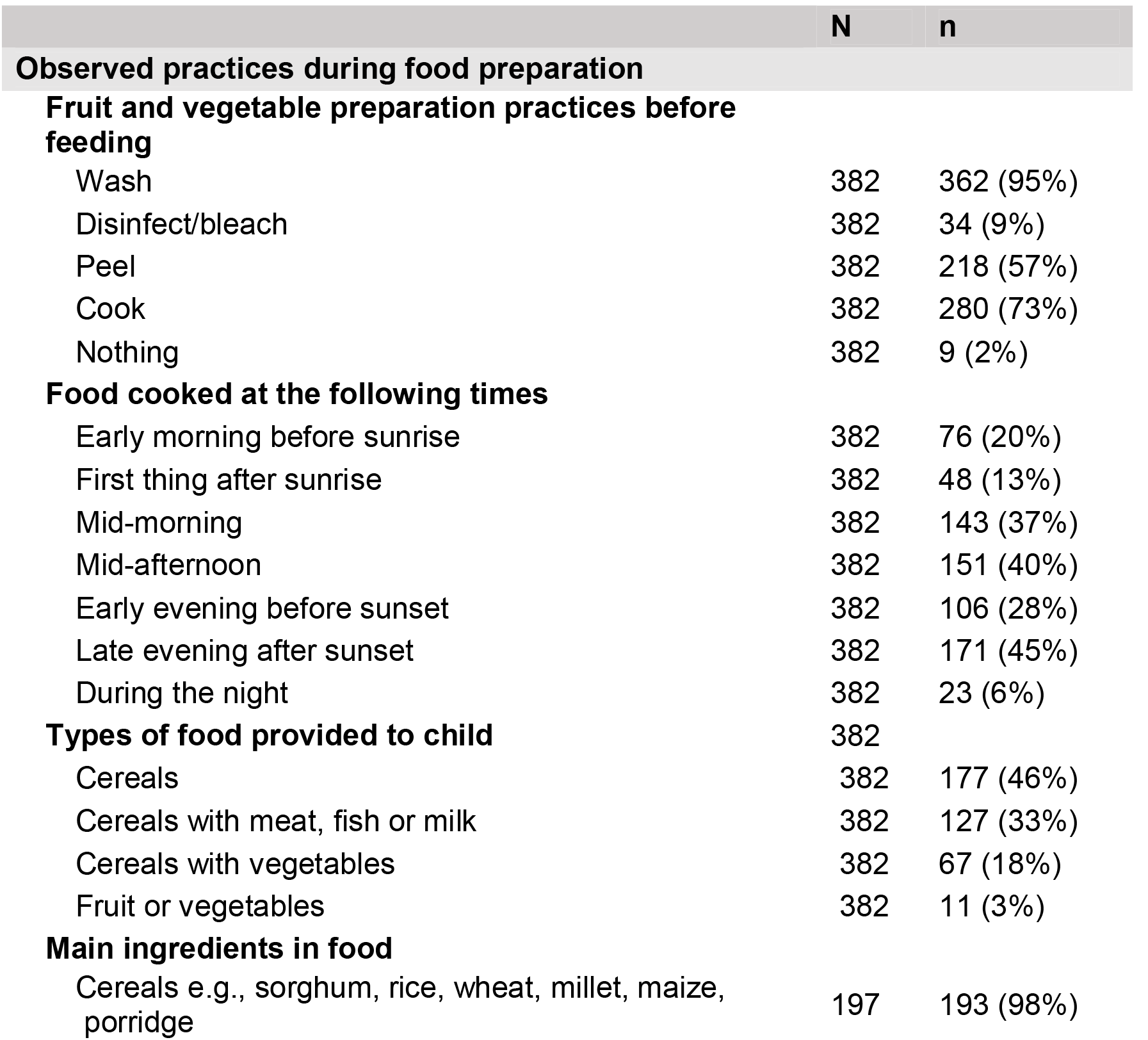
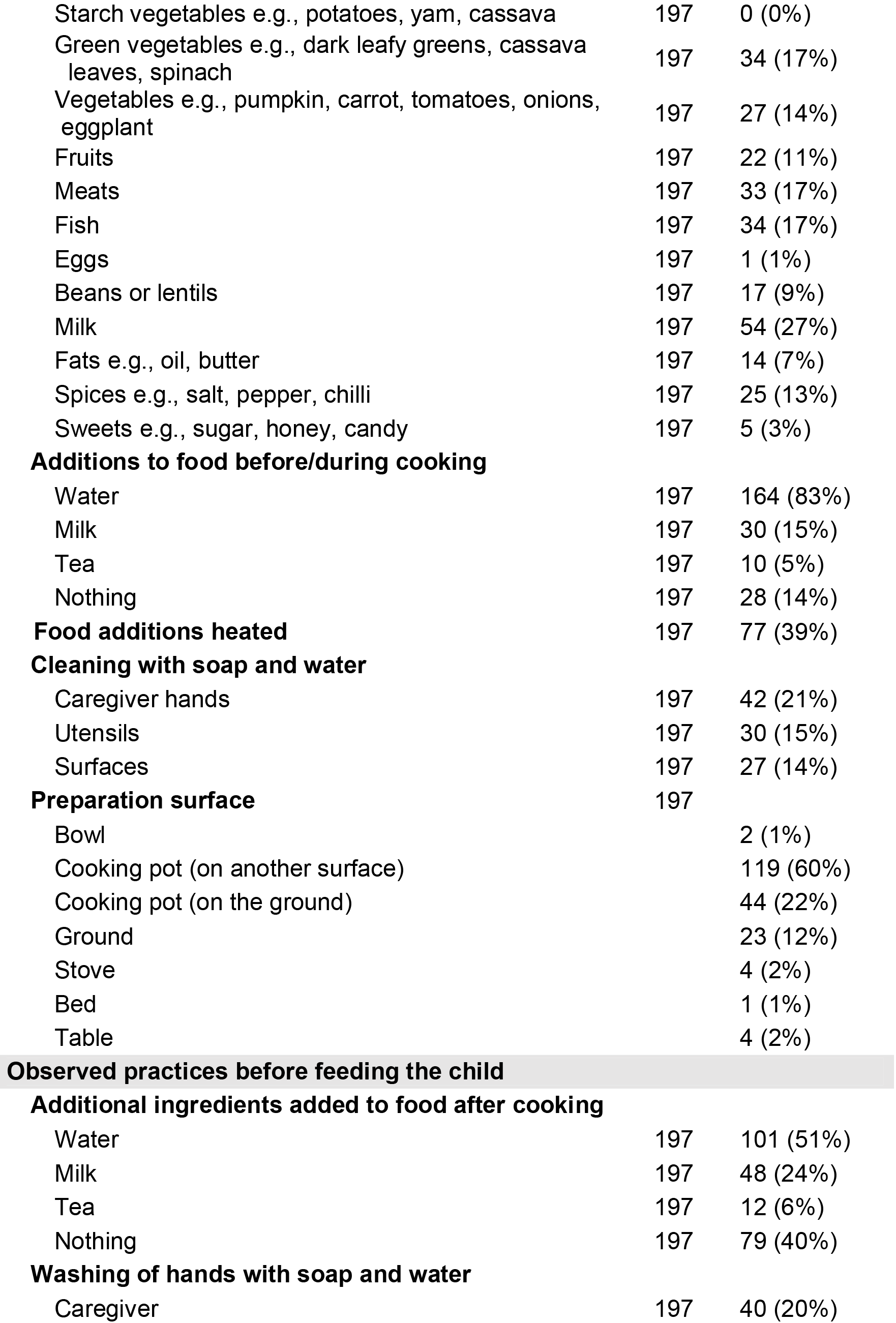
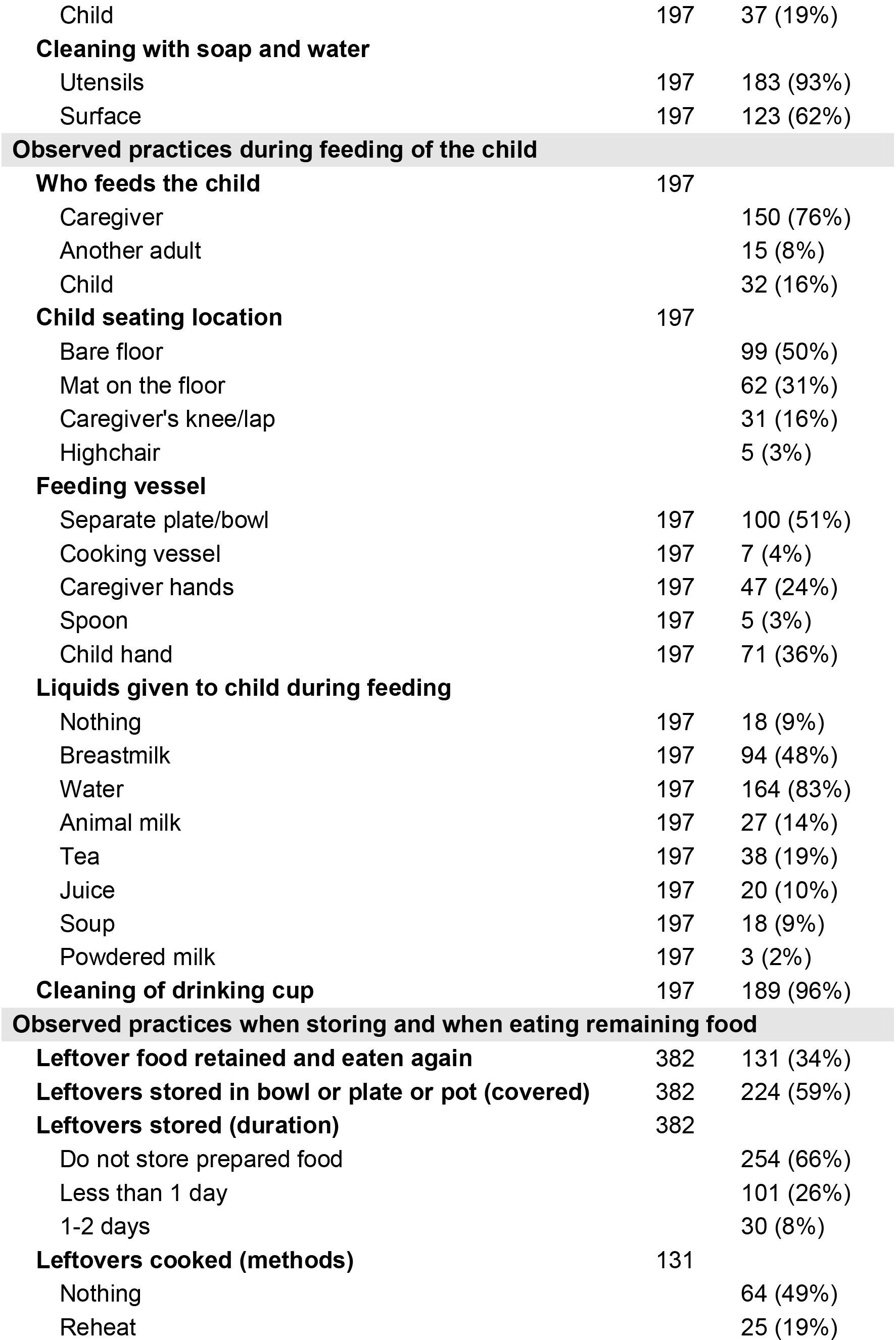
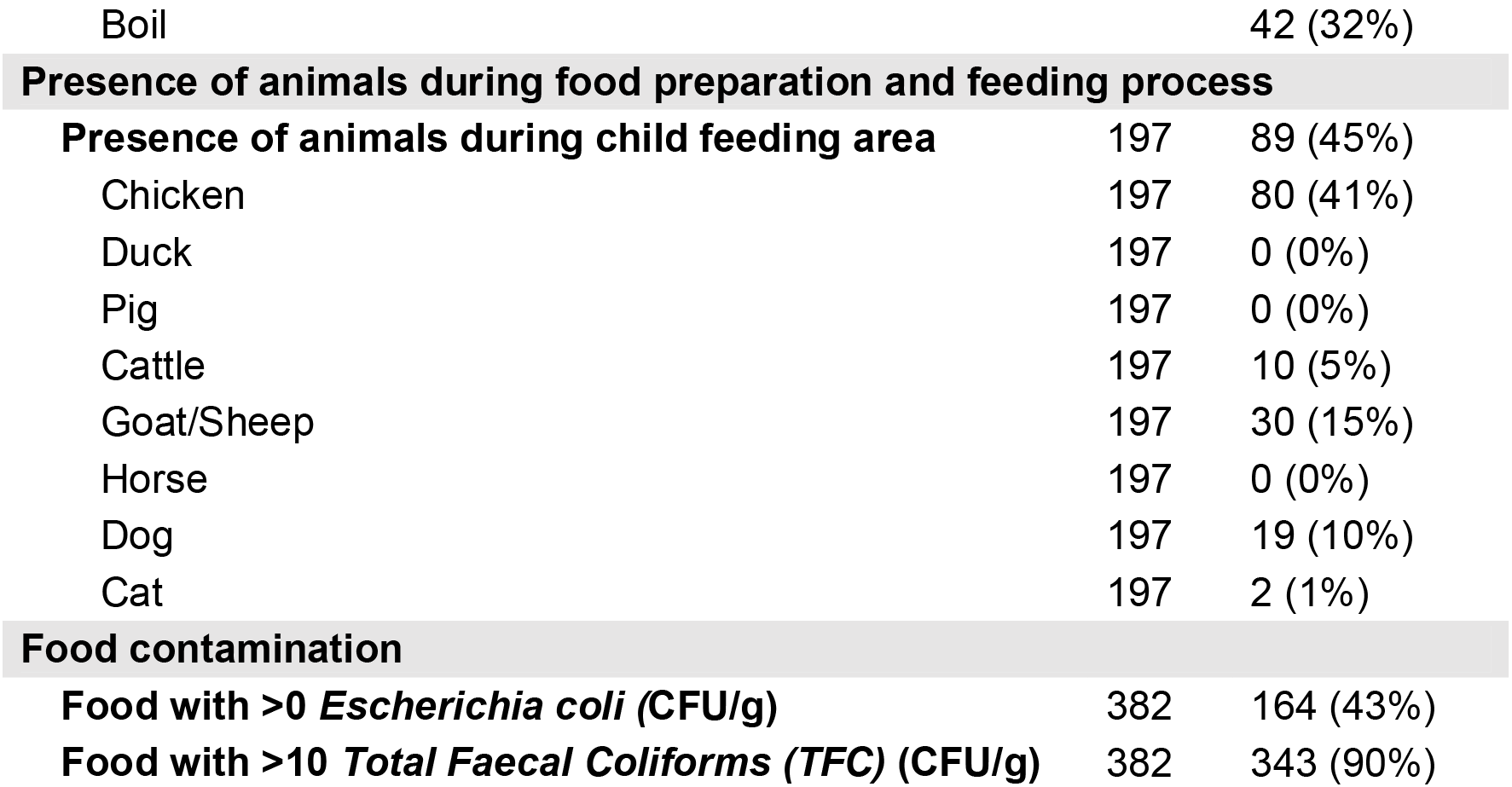
Food preparation, storage and feeding practices, including the prevalence of food contamination, in households of children recovered from severe acute malnutrition (SAM), Aweil East, South Sudan.

|  | N | n |
| --- | --- | --- |
| **Observed practices during food preparation** |  |  |
| **Fruit and vegetable preparation practices before feeding** |  |  |
| Wash | 382 | 362 (95%) |
| Disinfect/bleach | 382 | 34 (9%) |
| Peel | 382 | 218 (57%) |
| Cook | 382 | 280 (73%) |
| Nothing | 382 | 9 (2%) |
| **Food cooked at the following times** |  |  |
| Early morning before sunrise | 382 | 76 (20%) |
| First thing after sunrise | 382 | 48 (13%) |
| Mid-morning | 382 | 143 (37%) |
| Mid-afternoon | 382 | 151 (40%) |
| Early evening before sunset | 382 | 106 (28%) |
| Late evening after sunset | 382 | 171 (45%) |
| During the night | 382 | 23 (6%) |
| **Types of food provided to child** | 382 |  |
| Cereals | 382 | 177 (46%) |
| Cereals with meat, fish or milk | 382 | 127 (33%) |
| Cereals with vegetables | 382 | 67 (18%) |
| Fruit or vegetables | 382 | 11 (3%) |
| **Main ingredients in food** |  |  |
| Cereals e.g., sorghum, rice, wheat, millet, maize, porridge | 197 | 193 (98%) |
| Starch vegetables e.g., potatoes, yam, cassava | 197 | 0 (0%) |
| Green vegetables e.g., dark leafy greens, cassava leaves, spinach | 197 | 34 (17%) |
| Vegetables e.g., pumpkin, carrot, tomatoes, onions, eggplant | 197 | 27 (14%) |
| Fruits | 197 | 22 (11%) |
| Meats | 197 | 33 (17%) |
| Fish | 197 | 34 (17%) |
| Eggs | 197 | 1 (1%) |
| Beans or lentils | 197 | 17 (9%) |
| Milk | 197 | 54 (27%) |
| Fats e.g., oil, butter | 197 | 14 (7%) |
| Spices e.g., salt, pepper, chilli | 197 | 25 (13%) |
| Sweets e.g., sugar, honey, candy | 197 | 5 (3%) |
| **Additions to food before/during cooking** |  |  |
| Water | 197 | 164 (83%) |
| Milk | 197 | 30 (15%) |
| Tea | 197 | 10 (5%) |
| Nothing | 197 | 28 (14%) |
| **Food additions heated** | 197 | 77 (39%) |
| **Cleaning with soap and water** |  |  |
| Caregiver hands | 197 | 42 (21%) |
| Utensils | 197 | 30 (15%) |
| Surfaces | 197 | 27 (14%) |
| **Preparation surface** | 197 |  |
| Bowl |  | 2 (1%) |
| Cooking pot (on another surface) |  | 119 (60%) |
| Cooking pot (on the ground) |  | 44 (22%) |
| Ground |  | 23 (12%) |
| Stove |  | 4 (2%) |
| Bed |  | 1 (1%) |
| Table |  | 4 (2%) |
| **Observed practices before feeding the child** |  |  |
| **Additional ingredients added to food after cooking** |  |  |
| Water | 197 | 101 (51%) |
| Milk | 197 | 48 (24%) |
| Tea | 197 | 12 (6%) |
| Nothing | 197 | 79 (40%) |
| **Washing of hands with soap and water** |  |  |
| Caregiver | 197 | 40 (20%) |

|  | **N** | **n** |
| --- | --- | --- |
| Child | 197 | 37 (19%) |
| **Cleaning with soap and water** |  |  |
| Utensils | 197 | 183 (93%) |
| Surface | 197 | 123 (62%) |
| **Observed practices during feeding of the child** |  |  |
| **Who feeds the child** | 197 |  |
| Caregiver |  | 150 (76%) |
| Another adult |  | 15 (8%) |
| Child |  | 32 (16%) |
| **Child seating location** | 197 |  |
| Bare floor |  | 99 (50%) |
| Mat on the floor |  | 62 (31%) |
| Caregiver's knee/lap |  | 31 (16%) |
| Highchair |  | 5 (3%) |
| **Feeding vessel** |  |  |
| Separate plate/bowl | 197 | 100 (51%) |
| Cooking vessel | 197 | 7 (4%) |
| Caregiver hands | 197 | 47 (24%) |
| Spoon | 197 | 5 (3%) |
| Child hand | 197 | 71 (36%) |
| **Liquids given to child during feeding** |  |  |
| Nothing | 197 | 18 (9%) |
| Breastmilk | 197 | 94 (48%) |
| Water | 197 | 164 (83%) |
| Animal milk | 197 | 27 (14%) |
| Tea | 197 | 38 (19%) |
| Juice | 197 | 20 (10%) |
| Soup | 197 | 18 (9%) |
| Powdered milk | 197 | 3 (2%) |
| **Cleaning of drinking cup** | 197 | 189 (96%) |
| **Observed practices when storing and when eating remaining food** |  |  |
| **Leftover food retained and eaten again** | 382 | 131 (34%) |
| **Leftovers stored in bowl or plate or pot (covered)** | 382 | 224 (59%) |
| **Leftovers stored (duration)** | 382 |  |
| Do not store prepared food |  | 254 (66%) |
| Less than 1 day |  | 101 (26%) |
| 1-2 days |  | 30 (8%) |
| **Leftovers cooked (methods)** | 131 |  |
| Nothing |  | 64 (49%) |
| Reheat |  | 25 (19%) |
| Boil |  | 42 (32%) |
| **Presence of animals during food preparation and feeding process** |  |  |
| **Presence of animals during child feeding area** | 197 | 89 (45%) |
| Chicken | 197 | 80 (41%) |
| Duck | 197 | 0 (0%) |
| Pig | 197 | 0 (0%) |
| Cattle | 197 | 10 (5%) |
| Goat/Sheep | 197 | 30 (15%) |
| Horse | 197 | 0 (0%) |
| Dog | 197 | 19 (10%) |
| Cat | 197 | 2 (1%) |
| **Food contamination** |  |  |
| **Food with >0 *Escherichia coli* (CFU/g)** | 382 | 164 (43%) |
| **Food with >10 *Total Faecal Coliforms* (TFC) (CFU/g)** | 382 | 343 (90%) |

### Prevalence and quantity of food contamination

Food samples included: (a) cereals, including asida (a sorghum wheat-based pap), bread, rice and porridge (n = 177), (b) cereals with meat, fish and milk (n = 127), (c) cereals with vegetables (n = 67) and (d) fruit/vegetables (n = 11).

Of the 382 samples tested, 164 samples were contaminated with >0 *E. coli* CFU/g (43%, 95% CI [38%, 48%]) and 343 samples were contaminated with >10 TFC CFU/g (90%, 95% CI [87%, 93%]). The proportion of samples contaminated varied across food types (Figure 1). Cereals with fruit and vegetables were contaminated the most (51%, 95% CI [39%, 63%]), followed by cereals with meat, fish or milk (50%, 95% CI [41%, 58%]), fruit and vegetables (45%, 95% CI [19%, 74%]), and lastly, cereals served without any accompaniments (35%, 95% CI [28%, 42%]).

**Figure 1.**
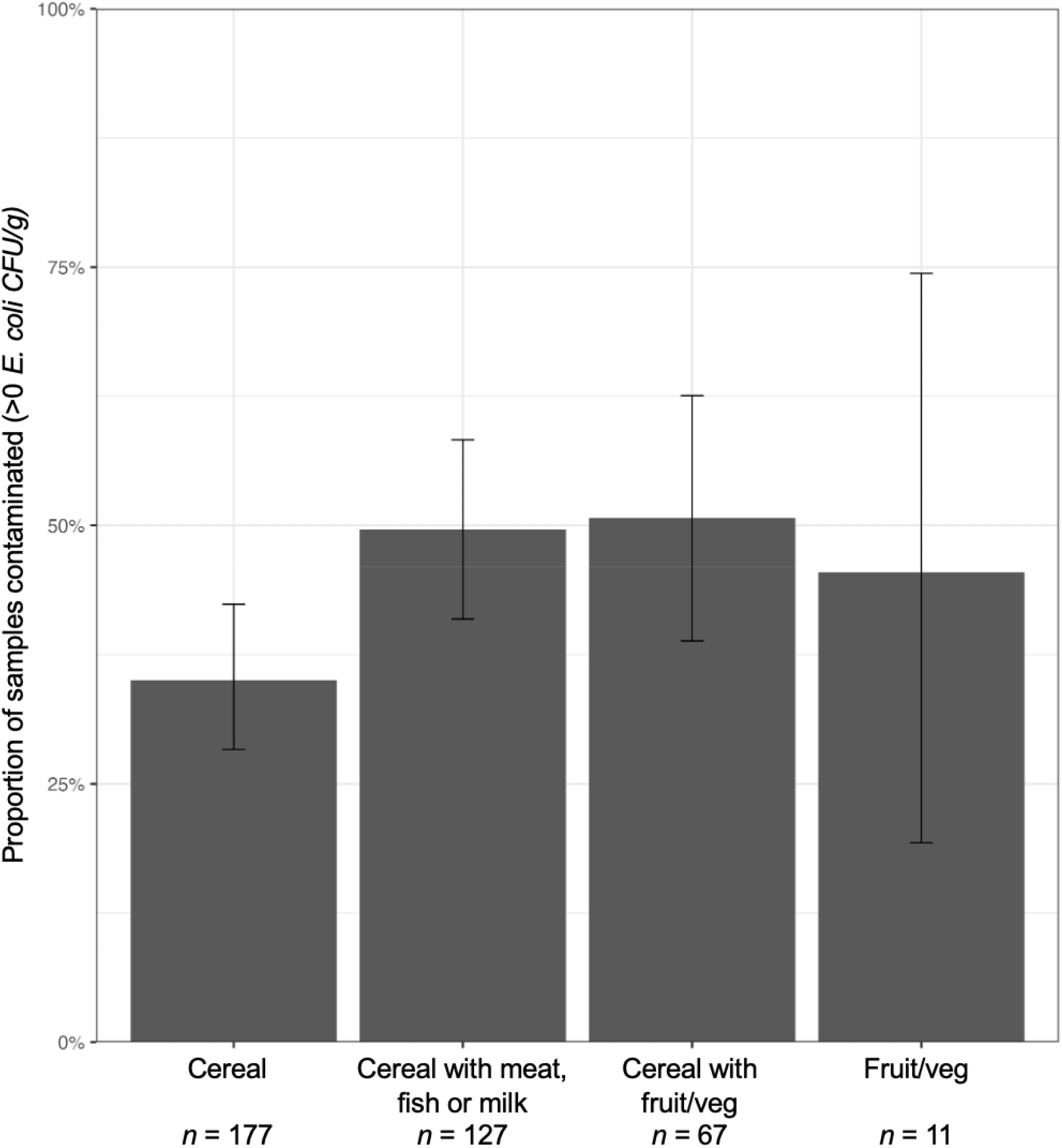
Proportion of child food samples contaminated with *E. coli* (>0 colony forming units [CFU] per gram [CFU/g] in households with children recovered from severe acute malnutrition (SAM) in Aweil East, South Sudan, by food type.

### HACCP analysis

The frequencies of observed preparation, feeding and storing practices of children’s food were reported for each of the four food categories (Table 3).

**Table 3.**
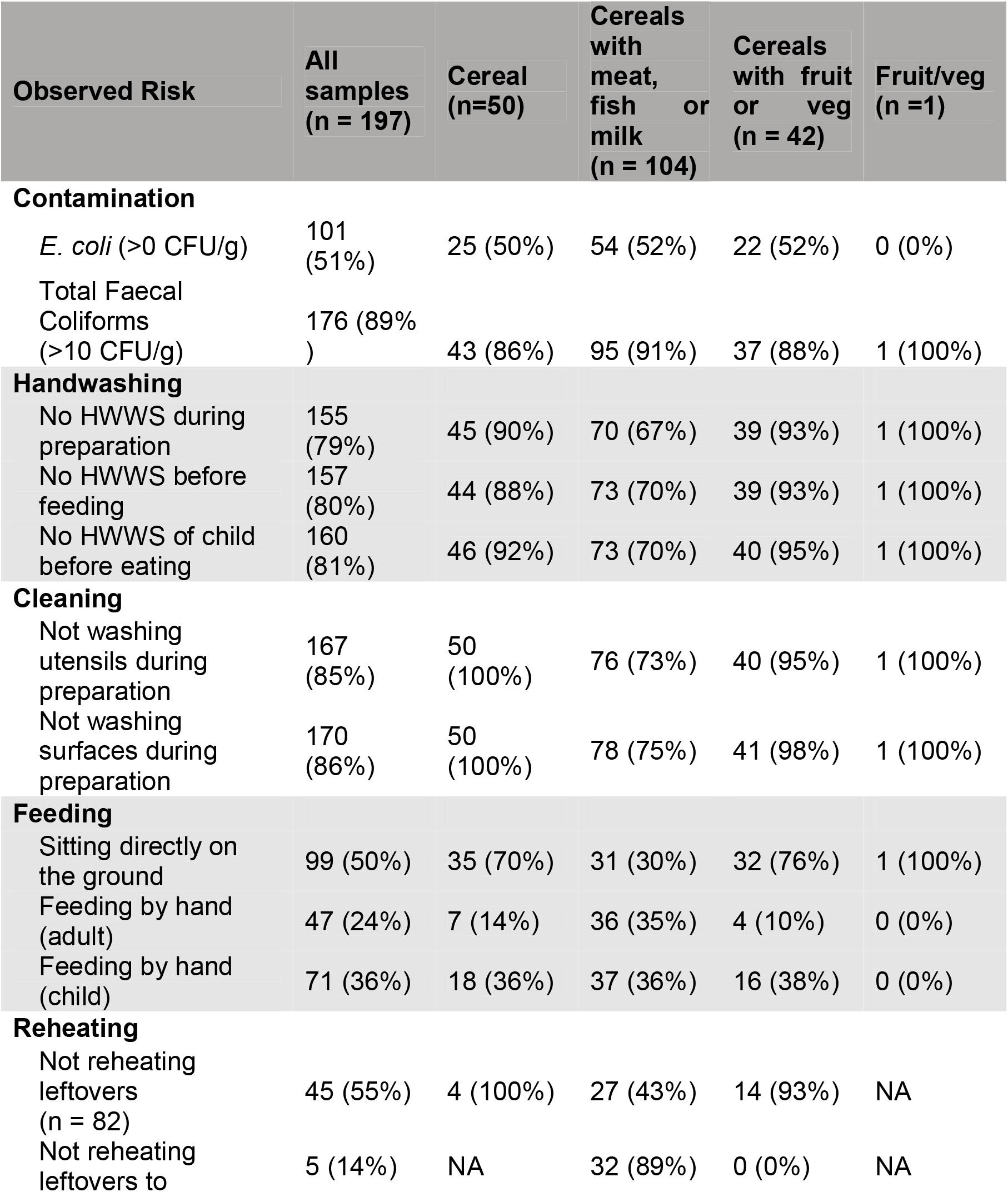
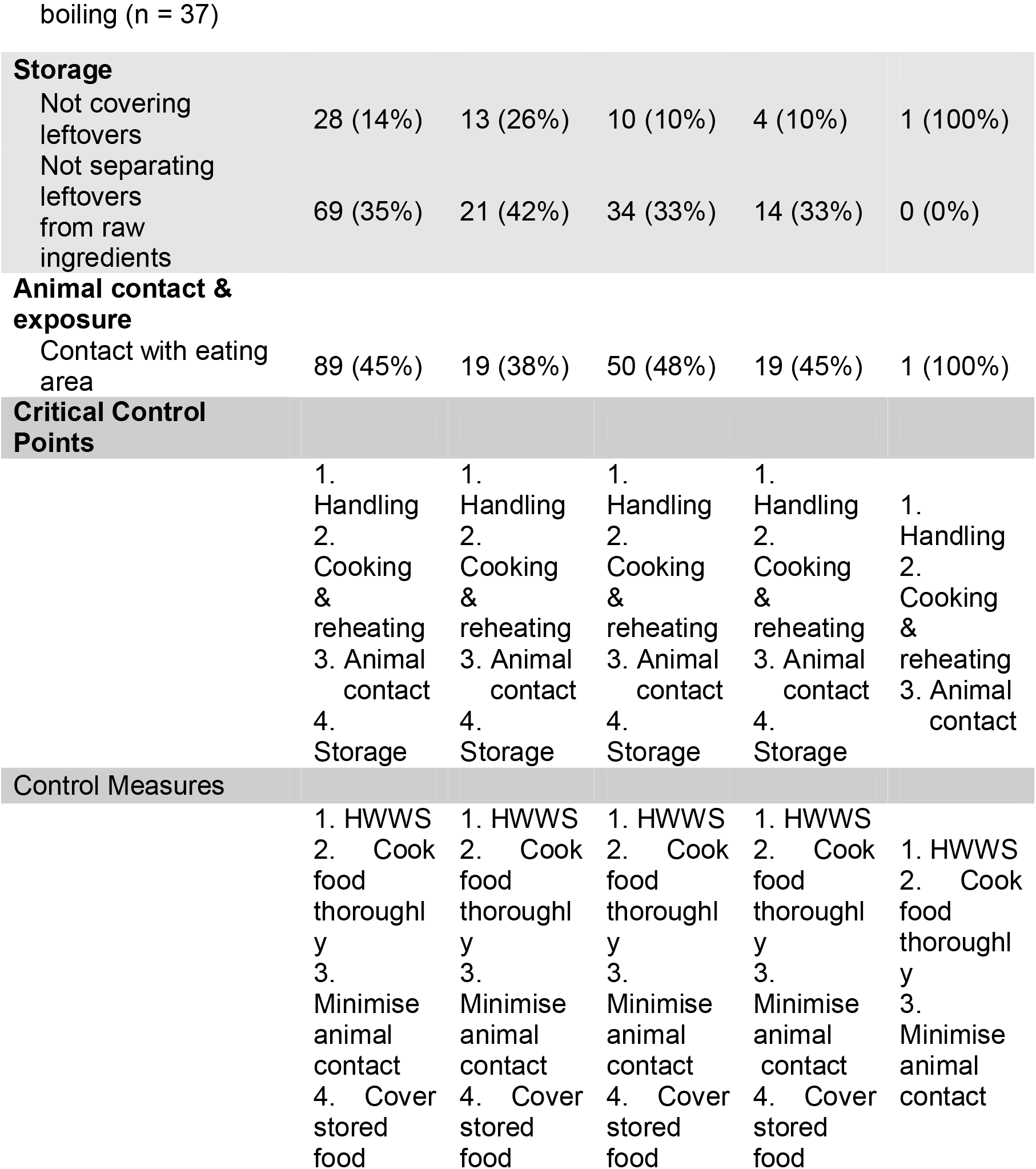
Observed food preparation and feeding practices, including critical control points and suggested control measures for food categories, among households of children recovered from severe acute malnutrition (SAM), in Aweil East, South Sudan.

Contamination with *E. coli* and TFCs was similar for all food categories. HWWS was rare by the caregiver before food preparation (21%), before feeding (20%), and of the child’s hands before eating (19%). Utensils (15%) and surfaces (14%) were not regularly washed before food preparation. Half (50%) of children were fed directly on the ground, with 24% of caregivers feeding the child with their hands, 3% feeding by spoon, or 36% of children feeding themselves by hand. When stored, 14% of food across all categories was not covered, which increased to 26% for cereals with meat, fish or milk. Of the stored food, 55% of meals were not reheated, with 35% of reheated food not being done to boiling. Many households also had animals in the child-feeding area (45%).

Food flow diagrams were produced for each food category based on the structured observations in 197 households; cereals with meat, fish or milk (n = 104) (Appendix B: Figure B1), cereal-based foods (n = 50) (Appendix B: Figure B2), cereals with fruit or vegetables (n = 42) (Appendix B: Figure B3), and fruit or vegetables (n = 1) (Appendix B: Figure B4). Each flow diagram was annotated with potential areas for hand, animal, water and utensil contamination, and CCPs. Specific food flow diagrams were combined to produce a general food flow diagram labelled with hypothesised areas of pathogen presence, growth and survival (Figure 2). The following control measures were suggested to counter the CCPs relevant for all food categories:

**Figure 2.**
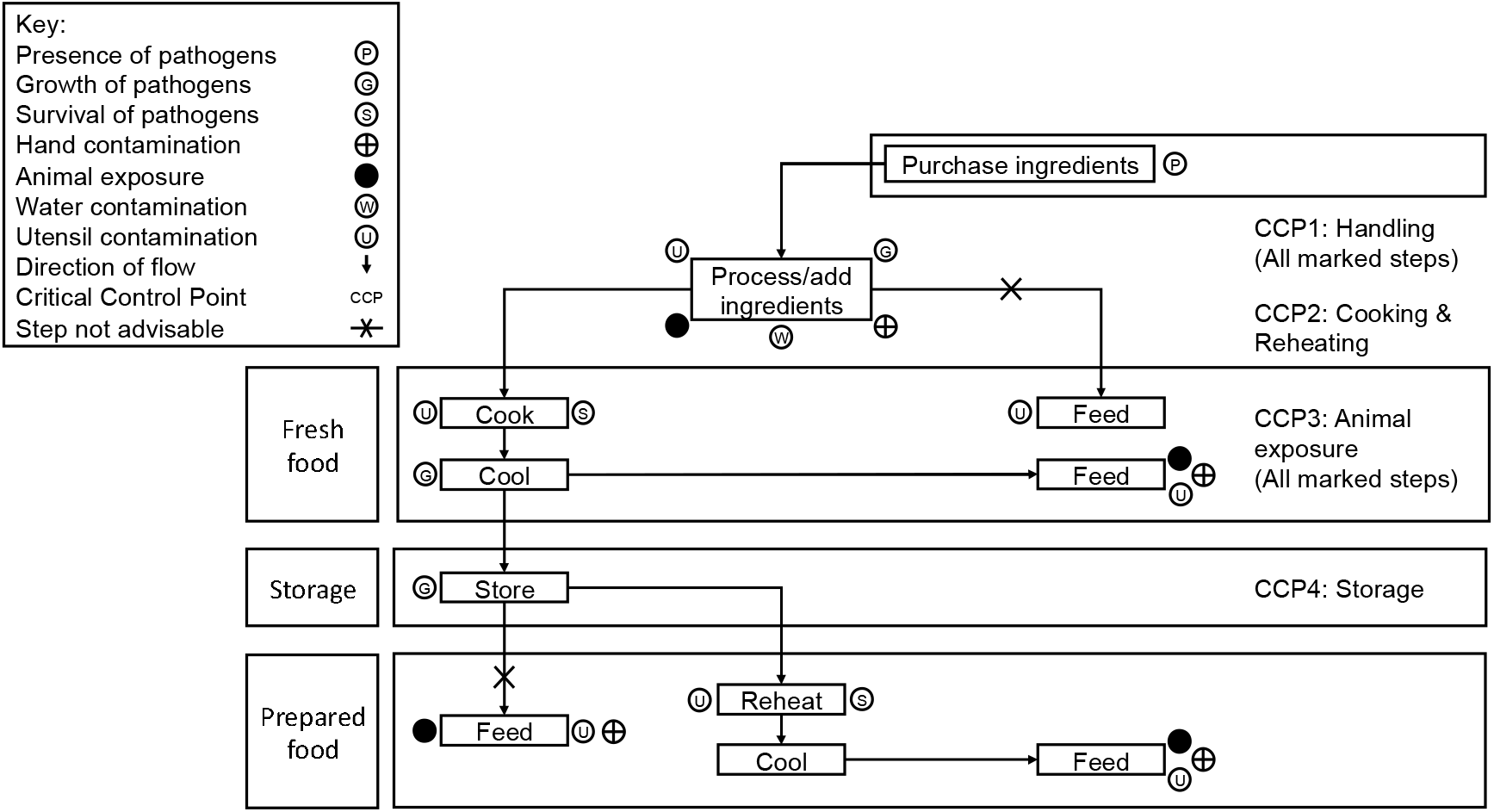
General food flow diagram for the preparation, feeding and storage of food, annotated with contamination risks and associated critical control points (CCPs), from households of children recovered from severe acute malnutrition (SAM), Aweil East, South Sudan.

1. HWWS throughout preparation and feeding and using clean utensils.
  - To limit CCP1: Handling (Figure 2).
  - HWWS of caregivers was low during food preparation (21%), before feeding (20%) and of the child (19%). Utensils were rarely used to feed the child (3%).
  - HWWS of both caregivers and children and using clean utensils are recommended to reduce food contamination.
2. Cook all food to boiling.
  - To limit CCP2: Cooking & Reheating (Figure 2).
  - Although leftovers were often reheated (55%), they were rarely done so to boiling (14%).
  - The reheating of prepared foods was often not observed, and when food was reheated, it was not always to boiling.
3. Avoid animal contact with the child’s eating area.
  - To limit CCP3: Animal exposure (Figure 2).
  - Various animal species were observed to come into contact with where the child is fed. 45% of households had at least one species come into contact with this area.
  - Avoiding animal contact with where the child is fed is suggested to reduce contamination of that area and food.
4. Store food in covered containers.
  - To limit CCP4: Storage (Figure 2).
  - Although food was more commonly covered (86%), not covering food (14%) was still observed and considered a risk.
  - Storing food in covered containers is justified to help reduce food contamination.

### Univariate regression for risk factors for *E. coli* (>0 CFU/g) and TFC contamination (>10 CFU/g) contamination of child foods

After the HACCP approach identified potential risk factors for food contamination, univariate regression models were run for potential risk factors and other covariates against the binary outcomes of *E. coli* (>0 CFU/g) and TFC contamination (>10 CFU/g). A total of eighteen variables were found to be significant (p < 0.1) when analysed against the binary outcome of *E. coli* contamination (>0 CFU/g) and eight variables for TFC contamination (>10 CFU/g). Univariate results can be found in Appendix C: Table C1.

### Multivariable regression for risk factors for *E. coli* (>0 CFU/g) and TFC contamination (>10 CFU/g) contamination of child foods

The multivariable regression model for food contamination >0 *E. coli* CFU/g included four variables (Table 4). Children feeding themselves (2.20 log [odds], 95% CI [1.16, 3.44]; p = <0.01) and animal presence in the child feeding area during mealtime (0.97 log [odds], 95% CI [0.28, 1.68]; p = <0.01) were found to be associated with increased *E. coli* contamination.

**Table 4.** Multivariable binomial regression model comprising four risk factors for child food contamination with >0 E. coli CFU/g in households of children recovered from severe acute malnutrition (SAM), Aweil East, South Sudan.

| Risk Factor | Coefficient | 2.5% CI | 97.5 % CI | *p* value |
| --- | --- | --- | --- | --- |
| Soap available - Yes (Ref: No) | -0.65 | -1.33 | 0.02 | 0.06 |
| Wash utensils before feeding - Yes (Ref: No) | 1.28 | -0.12 | 2.94 | 0.09 |
| Who is involved in feeding child (Ref: Primary caregiver) |  |  |  |  |
| Another adult | -0.36 | -1.63 | 0.84 | 0.56 |
| Child | 2.20 | 1.16 | 3.44 | <0.01 |
| Any animal was present in child feeding area during mealtime - Yes (Ref: No) | 0.97 | 0.28 | 1.68 | <0.01 |

Abbreviations: CI, confidence interval
Note: Adjusted for confounders: age, gender and socioeconomic status.

The multivariable regression model for food contamination >10 TFCs CFU/g comprised 1 variable (Table 5). Feeding the child with a spoon was found to have protective affects against TFC contamination (-4.17 log [odds], 95% CI [-7.81, -1.54]; p = <0.01).

**Table 5.** Multivariable binomial regression model comprising one risk factor for child food contamination with >10 *TFC* CFU/g in households of children recovered from severe acute malnutrition (SAM), Aweil East, South Sudan.

| Risk Factor | Coefficient | 2.5% CI | 97.5 % CI | *p* value |
| --- | --- | --- | --- | --- |
| Child fed with spoon - Yes (Ref: No) | -4.17 | -7.81 | -1.54 | <0.01 |

Abbreviations: CI, confidence interval
Note: Adjusted for confounders: age, gender and socioeconomic status.

## Discussion

This exploratory study assessed food contamination given to children aged 6-59 months that had recovered from SAM in Aweil East, South Sudan. To date, studies on food hygiene in similar settings or contexts are limited. The study combined a modified HACCP approach with qualitative structural observations and quantitative hazard data, collected through household surveys, to understand the factors contributing to the contamination of children’s food. In our study, 43% of the samples were contaminated with *E. coli* (>0 CFU/g), and 90% were contaminated with TFCs (>10 CFU/g). Four CCPs were identified during the preparation, feeding and storage of child food from the HACCP approach. These CCPs included the handling of raw and prepared food, cooking and reheating practices, animal contact during feeding, and storage of pre-prepared food. Child self-feeding and animal presence in the area during food preparation and feeding were risk factors for *E. coli* contamination. Feeding the child with a spoon was a protective factor against TFC contamination.

At the time of the study, there had been no similar studies into food contamination with *E. coli* in South Sudan, so direct comparisons are difficult to make. However, the 43% *E. coli* contamination levels (>0 *E. coli* CFU/g) in our study population are similar to that recorded in Bangladesh, with studies recording *E. coli* contamination of child foods between 39-46% (Islam et al., 2012; Muller-Hauser et al., 2022). Other studies have shown higher contamination levels in urban Mozambique (53% >10 enterococci CFU/g) (Bick et al., 2020) and rural Ethiopia (68% *E. coli* >0 CFU/g) (Gizaw et al., 2022). This study also found that 90% of food was contaminated with TFCs, slightly higher than the 84% of fresh and stored food contaminated with thermotolerant coliforms (TTC) (>10 TTC/g) found in peri-urban Mali (Touré et al., 2013). The contamination levels in South Sudan were in line with or higher than those found in other studies, and is especially concerning as expected levels of *E. coli* contamination in child food should be low or none (Hasell & Salter, 2003; Kennedy et al., 2011).

Other studies that carried out modified HACCP analysis of child foods observed similar food preparation, feeding and hygiene practices in their study populations, and drew similar conclusions to the CCPs in this study. Cooking, whether that is of fresh or pre-prepared food, food handling practices and storage were also identified in other studies (Bick et al., 2020; Gautam & Curtis, 2021; Islam et al., 2013; Manjang et al., 2018; Touré et al., 2011). Boiling of water or milk before being given to the child was also suggested as a CCP by other studies (Gautam & Curtis, 2021; Manjang et al., 2018). As this study did not assess the contamination of water or milk specifically, this was not found among our results but could be an area of further research. Our study also found animal contact with the food preparation and feeding area as a CCP whereas this was not identified in other studies (Bick et al., 2020).

The CCPs identified in this study aligned with the existing WHO “Five Keys to Safer Food” best practices for safer food, including 1) keeping hands, surfaces and equipment clean; 2) separating raw and cooked food; 3) cooking food thoroughly; 4) keeping food at safe temperatures and 5) using safe water (WHO, 2006). Whilst this study found no associations between the cooking and reheating or storage of food with contamination during the risk factor analysis, it did find evidence in the univariate analysis that the addition of meat, fish, milk, fruit or vegetables to cereal-based foods increased *E. coli* contamination. These foods are known to harbour *E. coli*, as well as other pathogens (Paudyal et al., 2017). The addition of these foods will increase the contamination of food and, if not cooked sufficiently to the recommended 70°C (WHO, 1996), remain contaminated. Access to cooking fuel can be a barrier to cooking and heating food sufficiently (Che, Zhu, & Wang, 2021), so appropriate solutions would have to be assessed. Similarly, this study found no association between safe food storage practices and contamination in the risk factor analysis, but serving prepared food was found to be associated with increased TFC contamination in the univariate analysis. Food storage practices, including temperature and duration, have been associated with contamination (Ercumen et al., 2017; Touré et al., 2011). Various studies have shown the effectiveness of covering food to reduce contamination (Ercumen et al., 2018; Parvez et al., 2017; Touré et al., 2011). Providing households with lidded containers could be an appropriate intervention to reduce contamination. In particular, air-vented containers mitigate concerns of spoiling, as suggested by a study in Bangladesh (Rahman et al., 2016).

Child self-feeding was found to be associated with increased *E. coli* contamination. A study in Bangladesh also identified the same risk factor (Islam et al., 2012) with several studies showing that children’s hands were typically contaminated with a high level of *E. coli* (Ercumen et al., 2017; Pickering et al., 2018). Children are likely to use their hands instead of utensils to eat, as well as touch surfaces including soil and the surrounding area (Davis et al., 2018). This creates a clear pathway for the contamination of children’s hands. If children’s hands are not cleaned before eating, as found in our study, there is a route for faecal contamination from their hands to food or directly to their mouths. HWWS of the child was rarely observed (19%), so improving the child’s hand hygiene may be an appropriate method to reduce food contamination. A trial in Nepal found that improving the handwashing of children and adults through activities including games, storytelling and local rallies could reduce food contamination risks (Gautam et al., 2017).

The presence of animals within the child feeding area during mealtime was found to be associated with increased food contamination with *E. coli.* Other studies in LMICs also found animals and the presence of animal faeces in the compound to be associated with food contamination (Barnes et al., 2018; Ercumen et al., 2017; Parvez et al., 2017). In this study, animals and their faeces were often not separated from the household, allowing for the contamination of soil and other surfaces (Penakalapati et al., 2017). Animals can host many foodborne enteric pathogens that can infect humans, including *Campylobacter*, *E. coli* and *Salmonella* (Heredia & García, 2018). Importantly, studies suggest that animal faecal contamination, especially from ruminant and avian species, is more prevalent than human contamination in the domestic environment, highlighting the role of animal faecal sources (Boehm et al., 2016; Schriewer et al., 2015). Studies have also observed that animal ownership increases *E. coli* contamination of soil and stored water (Ercumen et al., 2017; Navab-Daneshmand et al., 2018). This creates a clear pathway in which animal faeces can contaminate soil and water, which can then contaminate children’s hands and ultimately contaminate food. Improving animal husbandry practices such as keeping ruminant and avian species outside of the kitchen or eating areas may be an effective intervention to reducing child food contamination. This may include reducing the presence of animals within the compound or providing tools to dispose of faeces appropriately. However, local practices would need to be considered.

Feeding the child with a spoon was found to be a protective factor against TFC contamination. A very low proportion of households used a spoon to feed their child compared with other studies such as in Kenya, in which 92% of caregivers used a feeding utensil (spoon, bowl or cup) (Simiyu et al., 2022). A high proportion of households were recorded to have washed their utensils before feeding (93%), although whether this was with soap and water was not recorded. The use of clean utensils to prevent contamination aligns with the WHO’s best practices for safer food (WHO, 2006), helping to reduce the potential risk of hand contamination and may be an effective intervention in disrupting contamination from children’s hands.

The availability of soap within the household was found to have some protective effect (*p* = 0.06) against *E. coli* food contamination, which aligns with similar studies where soap had protective effects against food contamination (Bick et al., 2020; Davis et al., 2018; Islam et al., 2013). A meta-analysis found promotion of handwashing with soap reduced diarrhoea risk by 30% in LMICs (Wolf et al., 2022). However, the presence of soap does not equate to adequate use, especially in households that do not have sufficient quantity and quality of water to aid in handwashing (Abdullahi et al., 2020). This issue is also of concern for the effective washing of utensils, such as spoons. The households included in this study had low volumes of water stored in the house at the time of the survey which may imply that water for handwashing is a low priority. Providing soap to households may help to reduce the risk factors of child self-feeding and animal exposure, but alone may likely be ineffective without adequate handwashing facilities including access to and use of improved water sources.

## Limitations

As this is a cross-sectional study, the causality between risk factors and food contamination cannot be determined. The observations also likely excluded variables that contributed to food contamination such as cooking and storage temperatures and the presence of flies in the household.

As some of the data for this study were collected through structured observations, this may have led to reactivity through the Hawthorne effect, where participants modify an aspect of their behaviour in response to their awareness of being observed (Sedgwick & Greenwood, 2015). Furthermore, the observations by the reporter will likely have suffered from bias in their reporting, the enumerators may not have been able to observe all characteristics or there is the potential of “suppression” of data as they build a relationship with the observed participants.

Only taking one sample before feeding also limited our understanding of how contamination changes throughout the food preparation and feeding process. It would be useful to take repeated samples to pinpoint where contamination is introduced or reduced. HACCP analysis could be applied in more depth and detail if we had available contamination data and CCPs developed to a greater degree. The temperature and duration of cooking and storage could also have been recorded to help understand how this affects food contamination.

Using *E. coli* and TFCs as indicators for child food contamination provided only a limited understanding of the prevalence and diversity of other foodborne diseases in the child’s food. Understanding the individual pathogens present can help to determine their origins and possible routes of food contamination, and further in-depth analysis of food samples with multi-pathogen PCR analysis or other techniques is recommended.

## Conclusions

Our study found that over 40% of child food samples were contaminated, with child self-feeding and animal exposure being key risk factors for increased contamination. Our findings are consistent with other studies exploring child food contamination, particularly in low-income areas. This study helps to build on the understanding of child food contamination, especially in the context of children that have recovered from SAM - an area yet to be well researched. Developing our knowledge of the complex pathways in which food can become contaminated highlights that food hygiene interventions may complement more traditional WASH and infant and young child feeding (IYCF) interventions for children recovered from SAM. Simple control measures such as reducing animal contact in the household and during feeding; improving knowledge and practices around cooking; and, hygiene specific to the child may greatly reduce food contamination. This is of particular importance among children that have recovered from SAM and may help to protect this vulnerable population and reduce the risk of relapse to SAM.

## Data Availability

All data produced in the present study are available upon reasonable request to the authors

## Acknowledgements

We acknowledge the important contributions of all study participants and their families; the Ministries of Health in South Sudan; and the teams of dedicated staff who collected data for this study and provide treatment to malnourished children across all the study clinics.

## i. References

Abdullahi, L., Onyango, J. J., Mukiira, C., Wamicwe, J., Githiomi, R., Kariuki, D., . . . Mayieka, L. (2020). Community interventions in low-and middle-income countries to inform COVID-19 control implementation decisions in Kenya: A rapid systematic review. PLOS ONE, 15(12), e0242403. doi:10.1371/journal.pone.0242403

Action Against Hunger. (2021). Final report: Nutrition and mortality SMART survey in Aweil East County, NBeG State, South Sudan. Retrieved from Action Against Hunger, South Sudan.

Adegoke, O., Arif, S., Bahwere, P., Harb, J., Hug, J., Jasper, P., . . . Visram, A. (2021). Incidence of severe acute malnutrition after treatment: A prospective matched cohort study in Sokoto, Nigeria. Maternal & Child Nutrition, 17(1), e13070. doi:10.1111/mcn.13070

Ashraf, H., Alam, N. H., Chisti, M. J., Mahmud, S. R., Hossain, M. I., Ahmed, T., . . . Gyr, N. (2012). A follow-up experience of 6 months after treatment of children with severe acute malnutrition in Dhaka, Bangladesh. Journal of tropical pediatrics, 58(4), 253–257. doi:10.1093/tropej/fmr083

Barnes, A. N., Anderson, J. D., Mumma, J., Mahmud, Z. H., & Cumming, O. (2018). The association between domestic animal presence and ownership and household drinking water contamination among peri-urban communities of Kisumu, Kenya. PLOS ONE, 13(6), e0197587. doi:10.1371/journal.pone.0197587

Bick, S., Perieres, L., D’Mello-Guyett, L., Baker, K. K., Brown, J., Muneme, B., . . . Cumming, O. (2020). Risk factors for child food contamination in low-income neighbourhoods of Maputo, Mozambique: An exploratory, cross-sectional study. Maternal & Child Nutrition, 16(4), e12991. doi:10.1111/mcn.12991

Boehm, A. B., Wang, D., Ercumen, A., Shea, M., Harris, A. R., Shanks, O. C., . . . Pickering, A. J. (2016). Occurrence of host-associated fecal markers on child hands, household soil, and drinking water in rural Bangladeshi households. Environmental Science & Technology Letters, 3(11), 393–398. doi:10.1021/acs.estlett.6b00382

Che, X., Zhu, B., & Wang, P. (2021). Assessing global energy poverty: An integrated approach. Energy Policy, 149, 112099. doi:10.1016/j.enpol.2020.112099

Cichon, B., Fabiansen, C., Yaméogo, C. W., Rytter, M. J. H., Ritz, C., Briend, A., . . . Friis, H. (2016). Children with moderate acute malnutrition have inflammation not explained by maternal reports of illness and clinical symptoms: a cross-sectional study in Burkina Faso. BMC Nutrition, 2(1), 1–10. doi:10.1186/s40795-016-0096-0

Da Silva, N., Taniwaki, M. H., Junqueira, V. C., Silveira, N., Okazaki, M. M., & Gomes, R. A. R. (2018). Microbiological examination methods of food and water: a laboratory manual (2nd ed.). London: CRC Press.

Davis, E., Cumming, O., Aseyo, R. E., Muganda, D. N., Baker, K. K., Mumma, J., & Dreibelbis, R. (2018). Oral Contact events and caregiver hand hygiene: Implications for fecal-oral exposure to enteric pathogens among infants 3-9 months living in informal, peri-urban communities in Kisumu, Kenya. International Journal of Environmental Research and Public Health, 15(2), 192. doi:10.3390/ijerph15020192

Ercumen, A., Pickering, A. J., Kwong, L. H., Arnold, B. F., Parvez, S. M., Alam, M., . . . Colford, J. M., Jr. (2017). Animal feces contribute to domestic fecal contamination: evidence from E. coli measured in water, hands, food, flies, and soil in Bangladesh. Environmental Science & Technology, 51(15), 8725–8734. doi:10.1021/acs.est.7b01710

Ercumen, A., Pickering, A. J., Kwong, L. H., Mertens, A., Arnold, B. F., Benjamin-Chung, J., . . . Colford, J. M., Jr. (2018). Do sanitation improvements reduce fecal contamination of water, hands, food, soil, and flies? Evidence from a cluster-randomized controlled trial in rural Bangladesh. Environmental Science & Technology, 52(21), 12089–12097. doi:10.1021/acs.est.8b02988

Food and Agriculture Organization, & World Health Organization. (2017). Food Handlers Manual. Instructor. Retrieved from https://dx.doi.org/10.37774/9789275119020

Food and Agriculture Organization & World Health Organization Codex Alimentarius Commission. (1997). Hazard analysis and critical control point (HACCP) system and guidelines for its application. Annex to CAC/RCP 1-1969, Rev. 3. In Codex Alimentarius: Food hygiene basic texts – second edition (2001). Retrieved from https://www.fao.org/3/y1579e/y1579e03.htm#fn1

Gautam, O. P., & Curtis, V. (2021). Food hygiene practices of rural women and microbial risk for children: formative research in Nepal. The American Journal of Tropical Medicine and Hygiene, 105(5), 1383–1395. doi:10.4269/ajtmh.20-0574

Gautam, O. P., Schmidt, W. P., Cairncross, S., Cavill, S., & Curtis, V. (2017). Trial of a novel intervention to improve multiple food hygiene behaviors in Nepal. The American Journal of Tropical Medicine and Hygiene, 96(6), 1415–1426. doi:10.4269/ajtmh.16-0526

Gizaw, Z., Yalew, A. W., Bitew, B. D., Lee, J., & Bisesi, M. (2022). Fecal indicator bacteria along multiple environmental exposure pathways (water, food, and soil) and intestinal parasites among children in the rural northwest Ethiopia. BMC Gastroenterology, 22(1), 84. doi:10.1186/s12876-022-02174-4

Haas, C. N. (1996). How to average microbial densities to characterize risk. Water Research, 30(4), 1036–1038. doi:10.1016/0043-1354(95)00228-6

Hasell, S. K., & Salter, M. A. (2003). Review of the microbiological standards for foods. Food Control, 14(6), 391–398.

Havelaar, A. H., Kirk, M. D., Torgerson, P. R., Gibb, H. J., Hald, T., Lake, R. J., . . . Devleesschauwer, B. (2015). World Health Organization global estimates and regional comparisons of the burden of foodborne disease in 2010. PLoS Medicine, 12(12), e1001923. doi:10.1371/journal.pmed.1001923

Heredia, N., & García, S. (2018). Animals as sources of food-borne pathogens: A review. Animal Nutrition 4(3), 250–255. doi:10.1016/j.aninu.2018.04.006

Integrated Food Security Phase Classification. (2022). South Sudan: IPC acute food insecurity and acute malnutrition analysis *(February - July 2022)*. Retrieved from https://reliefweb.int/report/south-sudan/south-sudan-ipc-acute-food-insecurity-and-acute-malnutrition-analysis-february

Islam, M. A., Ahmed, T., Faruque, A. S., Rahman, S., Das, S. K., Ahmed, D., . . . Cravioto, A. (2012). Microbiological quality of complementary foods and its association with diarrhoeal morbidity and nutritional status of Bangladeshi children. European Journal of Clinical Nutrition, 66(11), 1242–1246. doi:10.1038/ejcn.2012.94

Islam, M. S., Mahmud, Z. H., Gope, P. S., Zaman, R. U., Hossain, Z., Islam, M. S., . . . Cairncross, S. (2013). Hygiene intervention reduces contamination of weaning food in Bangladesh. Tropical Medicine & International Health, 18(3), 250–258. doi:10.1111/tmi.12051

Kennedy, J., Gibney, S., Nolan, A., O’Brien, S., McMahon, M. A. S., McDowell, D., . . . Wall, P. G. (2011). Identification of critical points during domestic food preparation: an observational study. British Food Journal, 113(6), 766–783. doi:10.1108/00070701111140106

Khudri, M. M., & Chowdhury, F. (2013). Evaluation of socio-economic status of households and identifying key determinants of poverty in Bangladesh. European Journal of Social Sciences, 37(3), 377–387.

King, S., D’Mello-Guyett, L., Yakowenko, E., Riems, B., Gallandat, K., Mama Chabi, S., . . . Stobaugh, H. (2022). A multi-country, prospective cohort study to measure rate and risk of relapse among children recovered from severe acute malnutrition in Mali, Somalia, and South Sudan: a study protocol. BMC Nutrition, 8(1), 90. doi:10.1186/s40795-022-00576-x

Kotloff, K. L., Nataro, J. P., Blackwelder, W. C., Nasrin, D., Farag, T. H., Panchalingam, S., . . . Levine, M. M. (2013). Burden and aetiology of diarrhoeal disease in infants and young children in developing countries (the Global Enteric Multicenter Study, GEMS): a prospective, case-control study. The Lancet, 382(9888), 209–222. doi:10.1016/S0140-6736(13)60844-2

Manjang, B., Hemming, K., Bradley, C., Ensink, J., Martin, J. T., Sowe, J., . . . Manaseki-Holland, S. (2018). Promoting hygienic weaning food handling practices through a community-based programme: intervention implementation and baseline characteristics for a cluster randomised controlled trial in rural Gambia. BMJ Open, 8(8), e017573. doi:10.1136/bmjopen-2017-017573

Muller-Hauser, A. A., Sobhan, S., Huda, T. M. N., Waid, J. L., Wendt, A. S., Islam, M. A., . . . Gabrysch, S. (2022). Key food hygiene behaviors to reduce microbial contamination of complementary foods in rural Bangladesh. The American Journal of Tropical Medicine and Hygiene, 107(3), 709–719. doi:10.4269/ajtmh.21-0269

Navab-Daneshmand, T., Friedrich, M. N. D., Gachter, M., Montealegre, M. C., Mlambo, L. S., Nhiwatiwa, T., . . . Julian, T. R. (2018). Escherichia coli contamination across multiple environmental compartments (soil, hands, drinking water, and handwashing water) in urban Harare: correlations and risk factors. The American Journal of Tropical Medicine and Hygiene, 98(3), 803–813. doi:10.4269/ajtmh.17-0521

Ngure, F. M., Humphrey, J. H., Mbuya, M. N., Majo, F., Mutasa, K., Govha, M., . . . Stoltzfus, R. J. (2013). Formative research on hygiene behaviors and geophagy among infants and young children and implications of exposure to fecal bacteria. The American Journal of Tropical Medicine and Hygiene, 89(4), 709–716. doi:10.4269/ajtmh.12-0568

O’Sullivan, N. P., Lelijveld, N., Rutishauser-Perera, A., Kerac, M., & James, P. (2018). Follow-up between 6 and 24 months after discharge from treatment for severe acute malnutrition in children aged 6-59 months: A systematic review. PLOS ONE, 13(8), e0202053. doi:10.1371/journal.pone.0202053

Olofin, I., McDonald, C. M., Ezzati, M., Flaxman, S., Black, R. E., Fawzi, W. W., . . . Nutrition Impact Model, S. (2013). Associations of suboptimal growth with all-cause and cause-specific mortality in children under five years: a pooled analysis of ten prospective studies. PLOS ONE, 8(5), e64636. doi:10.1371/journal.pone.0064636

Paruch, A. M., & Mæhlum, T. (2012). Specific features of Escherichia coli that distinguish it from coliform and thermotolerant coliform bacteria and define it as the most accurate indicator of faecal contamination in the environment. Ecological Indicators, 23, 140–142. doi:10.1016/j.ecolind.2012.03.026

Parvez, S. M., Kwong, L., Rahman, M. J., Ercumen, A., Pickering, A. J., Ghosh, P. K., . . . Unicomb, L. (2017). Escherichia coli contamination of child complementary foods and association with domestic hygiene in rural Bangladesh. Tropical Medicine & International Health, 22(5), 547–557. doi:10.1111/tmi.12849

Paudyal, N., Anihouvi, V., Hounhouigan, J., Matsheka, M. I., Sekwati-Monang, B., Amoa-Awua, W., . . . Fang, W. (2017). Prevalence of foodborne pathogens in food from selected African countries - A meta-analysis. International Journal of Food Microbiology, 249, 35–43. doi:10.1016/j.ijfoodmicro.2017.03.002

Penakalapati, G., Swarthout, J., Delahoy, M. J., McAliley, L., Wodnik, B., Levy, K., & Freeman, M. C. (2017). Exposure to animal feces and human health: a systematic review and proposed research priorities. Environmental Science & Technology, 51(20), 11537–11552. doi:10.1021/acs.est.7b02811

Petri, W. A., Jr., Miller, M., Binder, H. J., Levine, M. M., Dillingham, R., & Guerrant, R. L. (2008). Enteric infections, diarrhea, and their impact on function and development. The Journal of Clinical Investigation 118(4), 1277–1290. doi:10.1172/JCI34005

Pickering, A. J., Ercumen, A., Arnold, B. F., Kwong, L. H., Parvez, S. M., Alam, M., . .. Luby, S. P. (2018). Fecal indicator bacteria along multiple environmental transmission pathways (water, hands, food, soil, flies) and subsequent child diarrhea in rural Bangladesh. Environmental Science & Technology, 52(14), 7928–7936. doi:10.1021/acs.est.8b00928

Rahman, M. J., Nizame, F. A., Nuruzzaman, M., Akand, F., Islam, M. A., Parvez, S. M., . . . Winch, P. J. (2016). Toward a scalable and sustainable intervention for complementary food safety. Food and Nutrition Bulletin, 37(2), 186–201. doi:10.1177/0379572116631641

Schaible, U. E., & Kaufmann, S. H. (2007). Malnutrition and infection: complex mechanisms and global impacts. PLoS Medicine, 4(5), e115. doi:10.1371/journal.pmed.0040115

Schriewer, A., Odagiri, M., Wuertz, S., Misra, P. R., Panigrahi, P., Clasen, T., & Jenkins, M. W. (2015). Human and animal fecal contamination of community water sources, stored drinking water and hands in rural India measured with validated microbial source tracking assays. The American Journal of Tropical Medicine and Hygiene, 93(3), 509–516. doi:10.4269/ajtmh.14-0824

Sedgwick, P., & Greenwood, N. (2015). Understanding the Hawthorne effect. BMJ, h4672. doi:10.1136/bmj.h4672

Simiyu, S., Aseyo, E., Anderson, J., Cumming, O., Baker, K. K., Dreibelbis, R., & Mumma, J. A. O. (2022). A mixed methods process evaluation of a food hygiene intervention in low-income informal neighbourhoods of Kisumu, Kenya. Maternal & Child Nutrition. doi:10.1007/s10995-022-03548-6

Stobaugh, H. C., Mayberry, A., McGrath, M., Bahwere, P., Zagre, N. M., Manary, M. J., . . . Lelijveld, N. (2019). Relapse after severe acute malnutrition: A systematic literature review and secondary data analysis. Maternal & Child Nutrition, 15(2), e12702. doi:10.1111/mcn.12702

Taulo, S., Wetlesen, A., Abrahamsen, R., Kululanga, G., Mkakosya, R., & Grimason, A. (2008). Microbiological hazard identification and exposure assessment of food prepared and served in rural households of Lungwena, Malawi. International Journal of Food Microbiology, 125(2), 111–116. doi:10.1016/j.ijfoodmicro.2008.02.025

Touré, O., Coulibaly, S., Arby, A., Maiga, F., & Cairncross, S. (2011). Improving microbiological food safety in peri-urban Mali; an experimental study. Food Control, 22(10), 1565–1572. doi:10.1016/j.foodcont.2011.03.012

Touré, O., Coulibaly, S., Arby, A., Maiga, F., & Cairncross, S. (2013). Piloting an intervention to improve microbiological food safety in Peri-Urban Mali. International Journal of Hygiene and Environmental Health, 216(2), 138–145. doi:10.1016/j.ijheh.2012.02.003

Tsai, K., Simiyu, S., Mumma, J., Aseyo, R. E., Cumming, O., Dreibelbis, R., & Baker, K. K. (2019). Enteric Pathogen Diversity in Infant Foods in Low-Income Neighborhoods of Kisumu, Kenya. International Journal of Environmental Research and Public Health, 16(3), 506. doi:10.3390/ijerph16030506

United Nations Children’s Fund. (2022). South Sudan Humanitarian Situation Report No. 12*: 1 January-31 December* 2022. Retrieved from https://reliefweb.int/report/south-sudan/unicef-south-sudan-humanitarian-situation-report-no-12-1-january-31-december-2022#:∼:text=In%202022%2C%208.9%20million%20people,with%20disabilities%20required%20humanitarian%20assistance.

United Nations Children’s Fund, World Health Organization, & World Bank. (2021). Levels and trends in child malnutrition: key findings of the 2021 edition of the joint child malnutrition estimates. Retrieved from https://www.who.int/publications/i/item/9789240025257

Vyas, S., & Kumaranayake, L. (2006). Constructing socio-economic status indices: how to use principal components analysis. Health Policy and Planning, 21(6), 459–468. doi:10.1093/heapol/czl029

Wolf, J., Hubbard, S., Brauer, M., Ambelu, A., Arnold, B. F., Bain, R., . . . Boisson, S. (2022). Effectiveness of interventions to improve drinking water, sanitation, and handwashing with soap on risk of diarrhoeal disease in children in low-income and middle-income settings: a systematic review and meta-analysis. The Lancet, 400(10345), 48–59. doi:10.1016/S0140-6736(22)00937-0

World Health Organization. (1996). Basic principles for the preparation of safe food for infants and young children. Retrieved from https://apps.who.int/iris/handle/10665/67833

World Health Organization. (2006). Five keys to safer food manual. Retrieved from https://www.who.int/publications/i/item/9789241594639

World Health Organization. (2013). Guideline: updates on the management of severe acute malnutrition in infants and children. Retrieved from https://www.who.int/publications/i/item/9789241506328

World Health Organization. (2015). WHO estimates of the global burden of foodborne diseases: foodborne disease burden epidemiology reference group 2007-2015 (9241565160). Retrieved from https://apps.who.int/iris/handle/10665/199350

World Health Organization, & United Nations Children’s Fund. (2023). The WHO/UNICEF joint monitoring programme estimates on WASH. Retrieved from https://washdata.org/

