## Appendices for "Risk factors for food contamination among children 6-59 months discharged from community management of acute malnutrition (CMAM) programmes for severe acute malnutrition (SAM) in Aweil East, South Sudan"

i. **Appendices**

Appendix A: Key data collection and analysis approaches

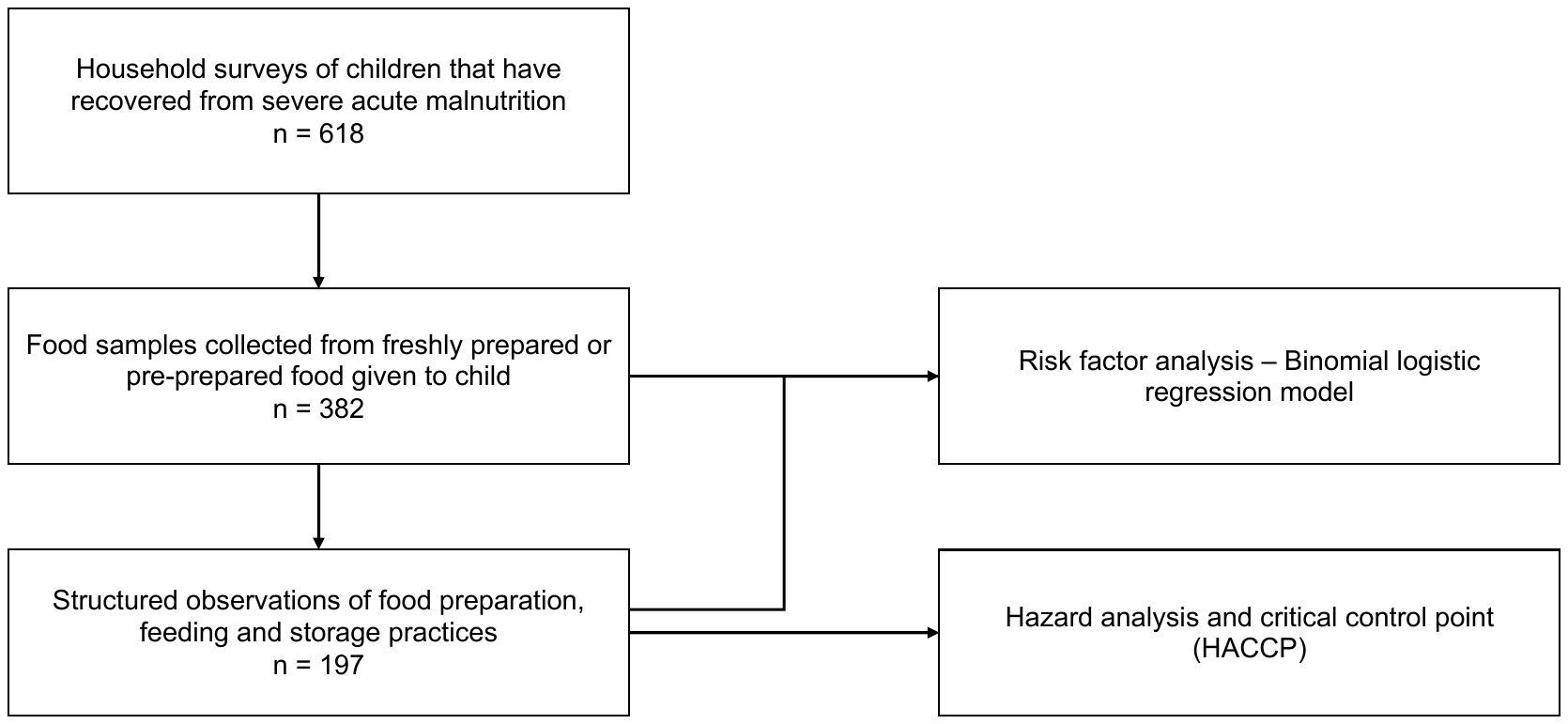
**Figure A1.** A participant flow diagram describing the data collection and analysis in a study to understand child food contamination in children recovered from severe acute malnutrition (SAM), Aweil East, South Sudan.

**Appendix B:** Food flow diagrams for four categories of child food

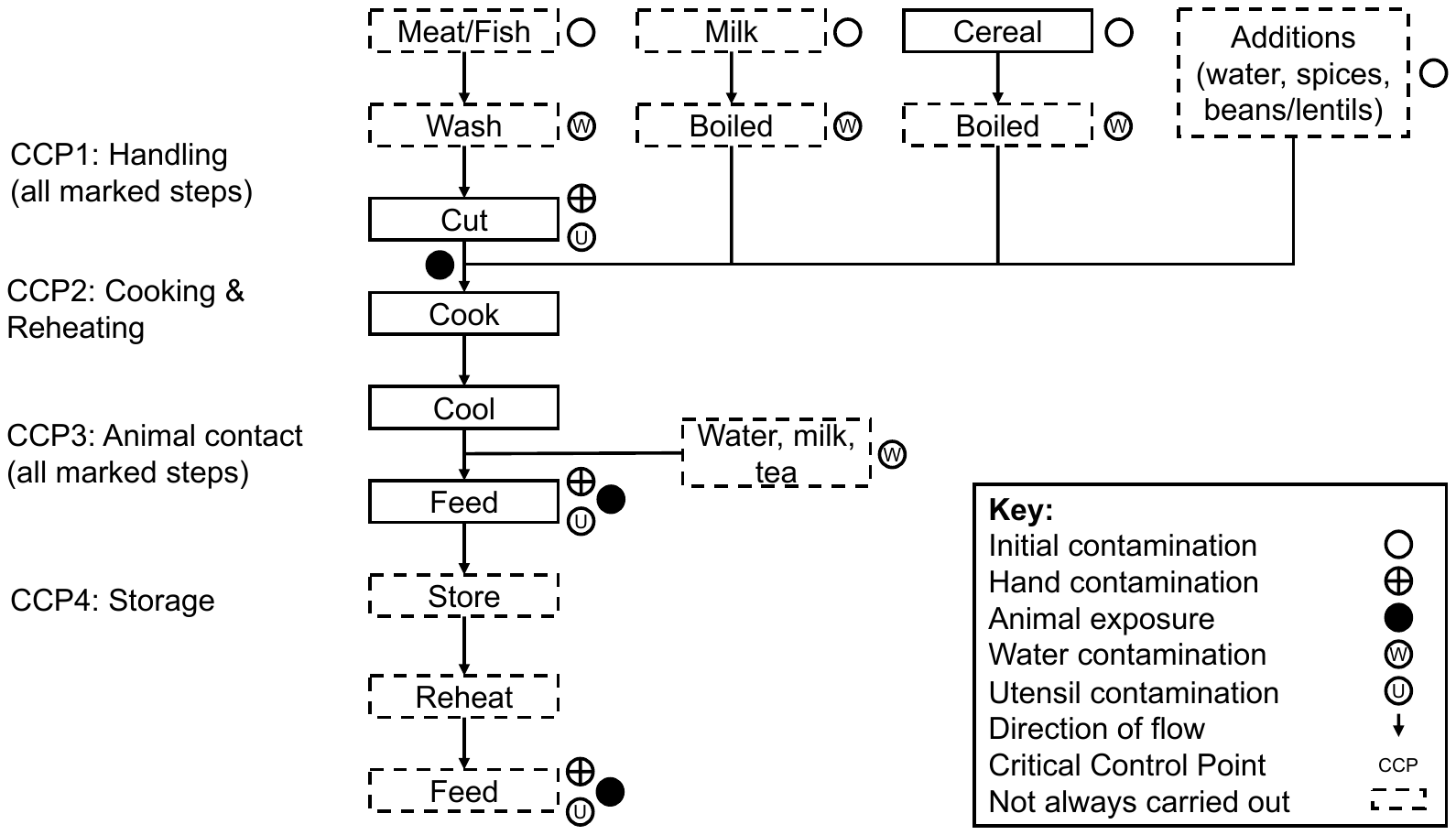

Note: These food-flow diagrams were constructed solely from structured observations.

**Figure B1.** Food flow diagram for the preparation, feeding and storage of meat, fish or milk, with cereal-based foods, annotated with contamination risks and critical control points (CCPs), in households of children recovered from severe acute malnutrition (SAM), Aweil East, South Sudan.

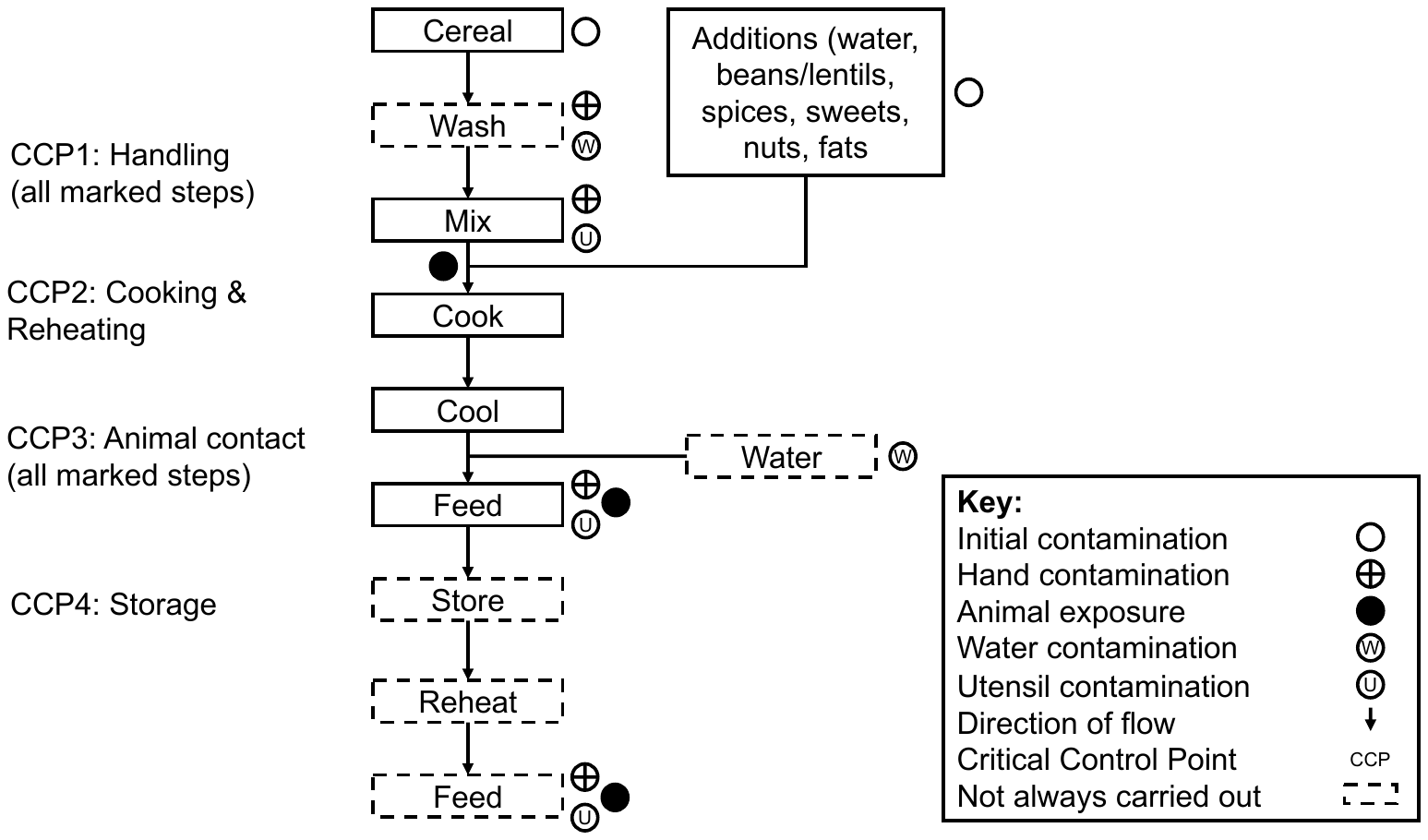

Note: These food-flow diagrams were constructed solely from structured observations.

**Figure B2.** Food flow diagram for the preparation, feeding and storage of cereal-based food (asida, porridge, rice and bread), annotated with contamination risks and critical control points (CCPs), in households of children recovered from severe acute malnutrition (SAM), Aweil East, South Sudan.

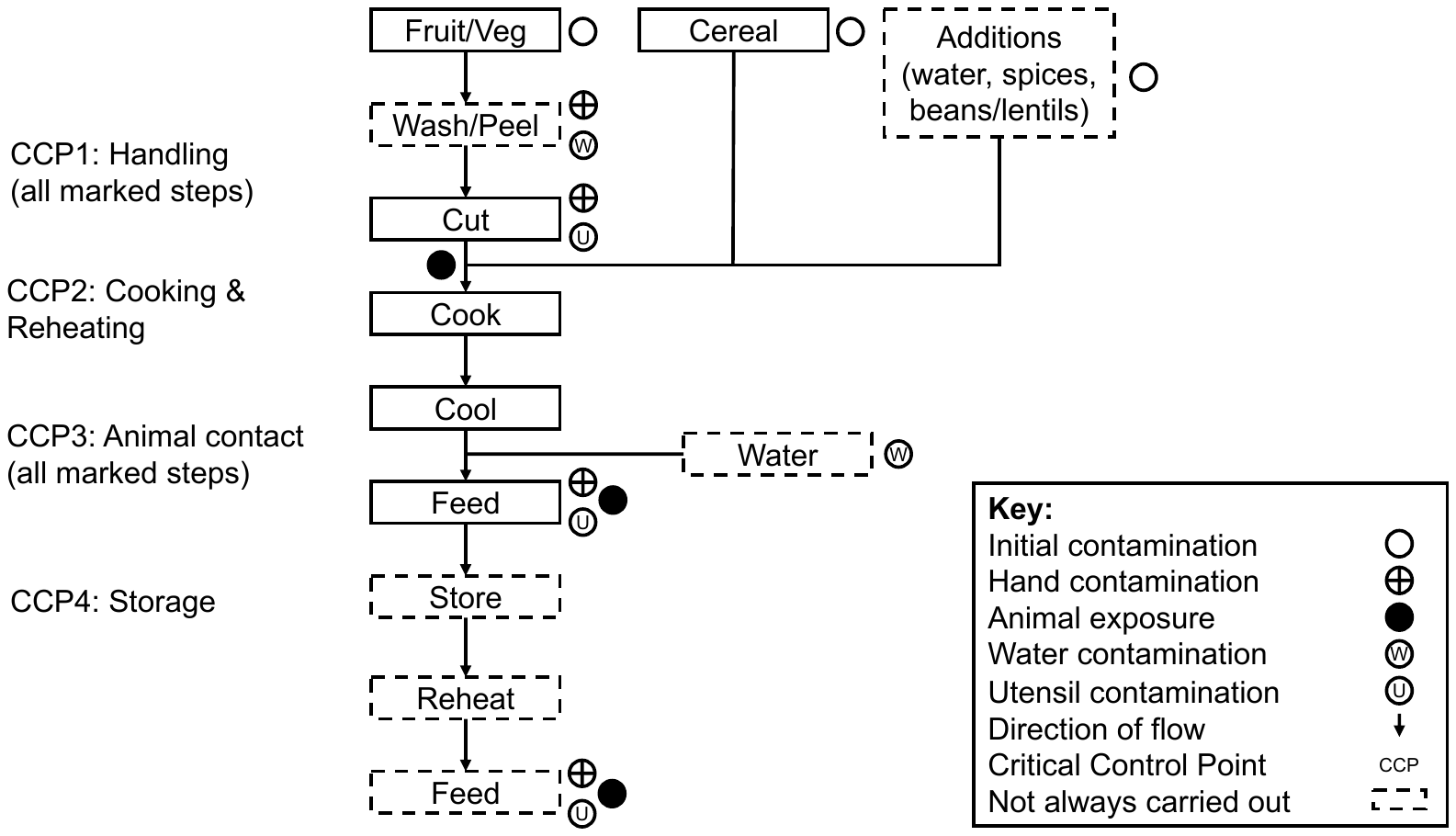

Note: These food-flow diagrams were constructed solely from structured observations.

**Figure B3.** Food flow diagram for the preparation, feeding and storage of cereal-based foods with fruit and/or vegetables, annotated with contamination risks and critical control points (CCPs), in households of children recovered from severe acute malnutrition (SAM), Aweil East, South Sudan.

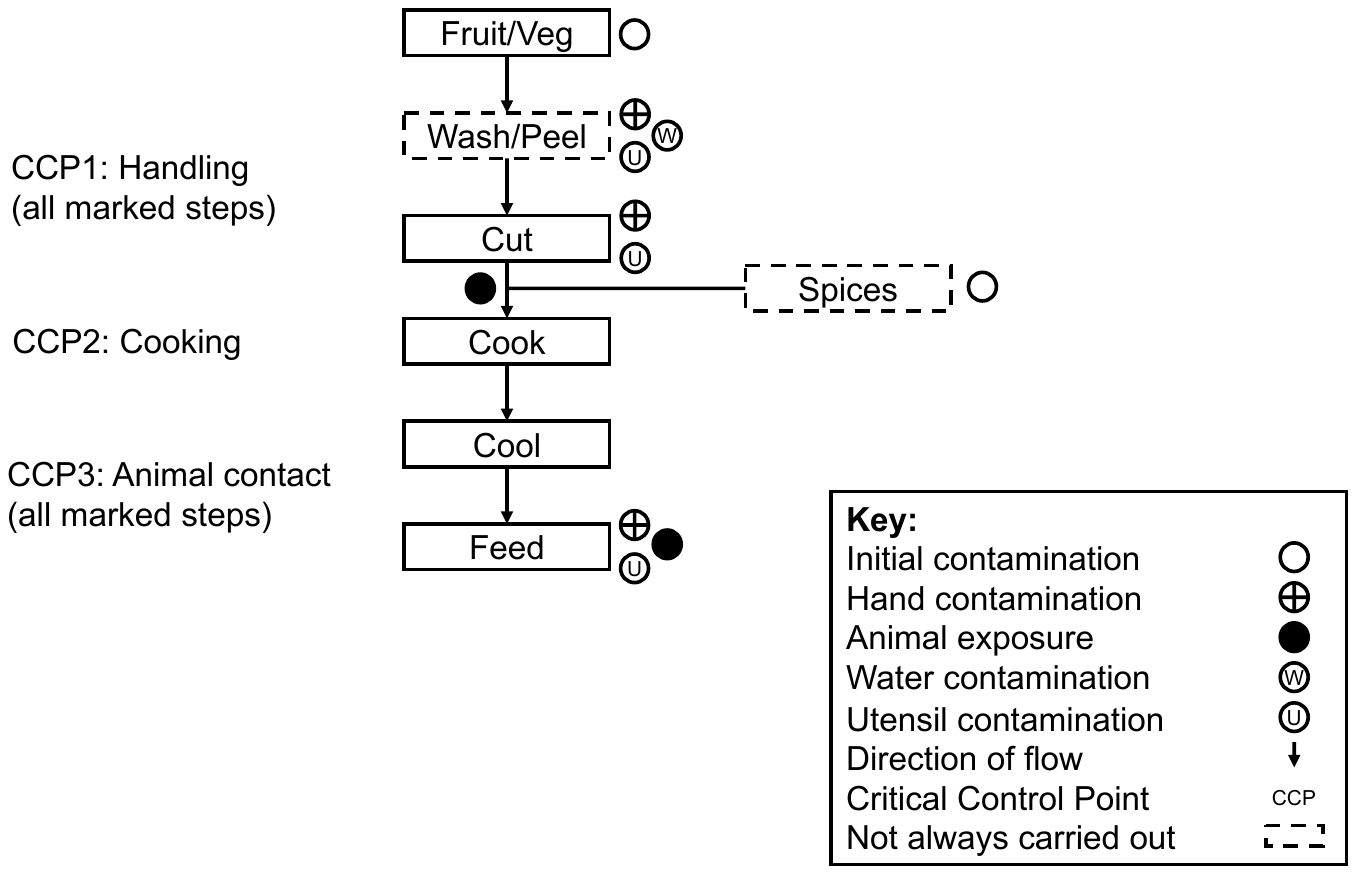

Note: These food-flow diagrams were constructed solely from structured observations.

**Figure B4.** Food flow diagram for the preparation, feeding and storage of fruit and/or vegetables, annotated with contamination risks and critical control points (CCPs), in households of children recovered from severe acute malnutrition (SAM), Aweil East, South Sudan.

**Appendix C:** Univariate analysis of household water, sanitation and hygiene (WASH) and food preparation and feeding exposures associated with *E. coli* and total faecal coliform (TFC) contamination

**Table C1.** Univariate binomial logistic regression model of 76 WASH and food preparation practices with *E. coli* (>0 CFU/g) and total faecal coliform (TFC) contamination (>10 CFU/g) in households of children recovered from severe acute malnutrition (SAM), Aweil East, South Sudan.

|  |  |  | **>0 *E. coli* CFU/g** | | | | | **>10 TFC CFU/g** | | | | |
| --- | --- | --- | --- | --- | --- | --- | --- | --- | --- | --- | --- | --- |
| **Variable** | **N** | **n (%)** | **Contamination** | **Coefficient** | **Lower 95% CI** | **Upper 95% CI** | ***p* value** | **Contamination** | **Coefficient** | **Lower 95% CI** | **Upper 95% CI** | ***p* value** |
| **Household exposure to animals (Ref: No)** |  |  |  |  |  |  |  |  |  |  |  |  |
| Any animal | 382 | 245 (64%) | 116 (47%) | 0.51 | 0.08 | 0.95 | 0.02 | 225 (92%) | 0.59 | -0.08 | 1.26 | 0.08 |
| Chickens | 382 | 207 (54%) | 104 (50%) | 0.66 | 0.25 | 1.08 | <0.05 | 189 (91%) | 0.36 | -0.31 | 1.03 | 0.29 |
| Ducks | 382 | 2 (1%) | 1 (50%) | 0.29 | -2.95 | 3.52 | 0.84 | 2 (100%) | Does not converge | | | |
| Pig | 382 | 1 (0%) | 1 (100%) | Does not converge | | | | 1 (100%) | Does not converge | | | |
| Goats/Sheep | 382 | 112 (29%) | 56 (50%) | 0.41 | -0.04 | 0.85 | 0.07 | 105 (94%) | 0.7 | -0.09 | 1.63 | 0.11 |
| Cattle | 382 | 64 (17%) | 31 (48%) | 0.27 | -0.27 | 0.81 | 0.33 | 61 (95%) | 0.95 | -0.11 | 2.4 | 0.12 |
| Horse | 382 | 2 (1%) | 0 (0%) | Does not converge | | | | 2 (100%) | Does not converge | | | |
| Dog | 382 | 51 (13%) | 22 (43%) | 0.01 | -0.59 | 0.60 | 0.98 | 48 (94%) | 0.67 | -0.4 | 2.12 | 0.28 |
| Cat | 382 | 12 (3%) | 6 (50%) | 0.29 | -0.89 | 1.47 | 0.62 | 11 (92%) | 0.23 | -1.45 | 3.15 | 0.83 |
| **Presence of animal faeces in compound - Yes (Ref: No)** | 382 | 136 (36%) | 64 (47%) | 0.26 | -0.16 | 0.68 | 0.23 | 116 (85%) | -0.72 | -1.4 | -0.05 | 0.03 |
| **Drinking water source** | 382 |  |  |  |  |  |  |  |  |  |  |  |
| Safely managed (improved source, accessible on premises, available when needed, free from contamination) |  | 300 (79%) | 127 (42%) | Ref. |  |  |  | 274 (91%) | Ref. |  |  |  |
| Basic (improved source, <30 min round trip) |  | 38 (10%) | 18 (47%) | 0.20 | -0.48 | 0.88 | 0.56 | 32 (84%) | -0.68 | -1.59 | 0.36 | 0.16 |
| Limited (improved source, >30 min round trip) |  | 29 (8%) | 12 (41%) | -0.04 | -0.83 | 0.73 | 0.92 | 25 (86%) | -0.52 | -1.56 | 0.75 | 0.36 |
| Unimproved (unprotected well/spring) |  | 15 (4%) | 7 (47%) | 0.18 | -0.90 | 1.22 | 0.74 | 12 (80%) | -0.97 | -2.19 | 0.55 | 0.15 |
| **Carry out water treatment practices - Yes (Ref: No)** | 382 | 91 (24%) | 38 (42%) | -0.06 | -0.54 | 0.41 | 0.80 | 79 (87%) | -0.40 | -1.10 | 0.36 | 0.28 |
| **Water storage practices (safely stored i.e., with cover) - Yes (Ref: No)** | 382 | 149 (39%) | 62 (42%) | -0.09 | -0.51 | 0.33 | 0.68 | 129 (87%) | -0.56 | -1.23 | 0.11 | 0.10 |
| **Experience of water scarcity (past 4 weeks)** | 382 |  |  |  |  |  |  |  |  |  |  |  |
| Never |  | 227 (59%) | 95 (42%) | Ref. |  |  |  | 204 (90%) | Ref. |  |  |  |
| Once or twice |  | 121 (32%) | 53 (44%) | 0.08 | -0.37 | 0.52 | 0.73 | 107 (88%) | -0.15 | -0.84 | 0.58 | 0.68 |
| 2 -10 times |  | 24 (6%) | 10 (42%) | -0.01 | -0.89 | 0.84 | 0.99 | 22 (92%) | 0.22 | -1.09 | 2.09 | 0.78 |
| More than 10 times |  | 10 (3%) | 6 (60%) | 0.73 | -0.55 | 2.12 | 0.27 | 10 (100%) | Does not converge | | | |
| **Use other sources of water - Yes (Ref: No)** | 382 | 231 (60%) | 103 (45%) | 0.17 | -0.24 | 0.59 | 0.42 | 206 (89%) | -0.17 | -0.88 | 0.50 | 0.62 |
| **Sanitation facility** | 382 |  |  |  |  |  |  |  |  |  |  |  |
| Limited (improved facility, shared with two or more households) |  | 35 (9%) | 13 (37%) | Ref. |  |  |  | 33 (94%) | Ref. |  |  |  |
| Unimproved (pit latrine without platform, hanging latrine  or bucket latrine) |  | 52 (14%) | 19 (37%) | -0.03 | -0.91 | 0.87 | 0.95 | 46 (88%) | -0.77 | -2.73 | 0.77 | 0.37 |
| Open defecation (disposal of faeces in environment  (e.g., fields, forests) |  | 295 (77%) | 132 (45%) | 0.32 | -0.40 | 1.06 | 0.39 | 264 (89%) | -0.66 | -2.51 | 0.60 | 0.38 |
| **Presence of human faeces in compound - Yes (Ref: No)** | 382 | 53 (14%) | 28 (53%) | 0.46 | -0.12 | 1.05 | 0.12 | 47 (89%) | -0.14 | -1.00 | 0.88 | 0.77 |
| **Handwashing facility** | 382 |  |  |  |  |  |  |  |  |  |  |  |
| Basic (available handwashing facility with soap and water at home) |  | 77 (20%) | 34 (44%) | Ref. |  |  |  | 69 (90%) | Ref. |  |  |  |
| Limited (available handwashing facility lacking soap and/or water) |  | 285 (75%) | 118 (41%) | -0.11 | -0.62 | 0.40 | 0.66 | 258 (91%) | 0.10 | -0.79 | 0.89 | 0.81 |
| No handwashing facility |  | 20 (5%) | 12 (60%) | 0.64 | -0.35 | 1.68 | 0.21 | 16 (80%) | -0.77 | -2.05 | 0.65 | 0.25 |
| **Soap available** | 382 | 181 (47%) | 68 (38%) | -0.42 | -0.83 | -0.01 | 0.04 | 161 (89%) | -0.17 | -0.84 | 0.49 | 0.61 |
| **Handwashing at more than 5 key times for HWWS - Yes (Ref: No)** | 382 | 107 (28%) | 53 (50%) | 0.37 | -0.08 | 0.82 | 0.10 | 99 (93%) | 0.45 | -0.31 | 1.33 | 0.27 |
| **Handwashing with soap and water - Yes (Ref: No)** | 382 | 148 (41%) | 65 (44%) | 0.27 | -0.15 | 0.68 | 0.21 | 142 (91%) | 0.23 | -0.44 | 0.94 | 0.51 |
| **Presence of garbage or waste in compound - Yes (Ref: No)** | 382 | 263 (69%) | 115 (44%) | 0.10 | -0.33 | 0.55 | 0.64 | 241 (92%) | 0.60 | -0.08 | 1.27 | 0.08 |
| **Food cooked at following times (Ref: No)** |  |  |  |  |  |  |  |  |  |  |  |  |
| Early morning before sunrise | 382 | 76 (20%) | 34 (45%) | 0.09 | -0.42 | 0.60 | 0.72 | 68 (89%) | -0.04 | -0.82 | 0.84 | 0.92 |
| First thing after sunrise | 382 | 48 (13%) | 20 (42%) | -0.06 | -0.68 | 0.55 | 0.85 | 40 (83%) | -0.67 | -1.48 | 0.23 | 0.12 |
| Mid-morning | 382 | 143 (37%) | 65 (45%) | 0.16 | -0.25 | 0.58 | 0.44 | 130 (91%) | 0.20 | -0.48 | 0.93 | 0.58 |
| Mid-afternoon | 382 | 151 (40%) | 61 (40%) | -0.17 | -0.59 | 0.24 | 0.42 | 138 (91%) | 0.30 | -0.39 | 1.03 | 0.40 |
| Early evening before sunset | 382 | 106 (28%) | 44 (42%) | -0.08 | -0.54 | 0.37 | 0.73 | 95 (90%) | -0.03 | -0.74 | 0.75 | 0.95 |
| Late evening after sunset | 382 | 171 (45%) | 73 (43%) | -0.02 | -0.43 | 0.39 | 0.93 | 158 (92%) | 0.54 | -0.15 | 1.26 | 0.13 |
| During the night | 382 | 23 (6%) | 11 (48%) | 0.21 | -0.65 | 1.06 | 0.63 | 22 (96%) | 0.96 | -0.65 | 3.86 | 0.36 |
| **Food categorisation** | 382 |  |  |  |  |  |  |  |  |  |  |  |
| Cereals |  | 177 (46%) | 62 (35%) | Ref. |  |  |  | 156 (88%) | Ref. |  |  |  |
| Cereals with meat, fish or milk |  | 127 (33%) | 63 (50%) | 0.60 | 0.14 | 1.07 | 0.01 | 116 (91%) | 0.35 | -0.40 | 1.15 | 0.37 |
| Cereals with vegetables |  | 67 (18%) | 34 (51%) | 0.65 | 0.08 | 1.22 | 0.03 | 61 (91%) | 0.31 | -0.59 | 1.35 | 0.52 |
| Fruit or vegetables |  | 11 (3%) | 5 (45%) | 0.44 | -0.84 | 1.67 | 0.49 | 10 (91%) | 0.30 | -1.43 | 3.23 | 0.78 |
| **Leftover food retained and eaten again - Yes (Ref: No)** | 382 | 131 (34%) | 100 (40%) | 0.36 | -0.07 | 0.79 | 0.10 | 118 (90%) | Does not converge | | | |
| **Food stored in covered container - Yes (Ref: No)** | 382 | 224 (59%) | 106 (47%) | 0.41 | -0.01 | 0.83 | 0.06 | 199 (89%) | Does not converge | | | |
| **Drinking water with >5 NTU - Yes (Ref: No)** | 382 | 143 (38%) | 59 (41%) | -0.10 | -0.52 | 0.32 | 0.65 | 130 (91%) | 0.17 | -0.52 | 0.90 | 0.64 |
| **Drinking water with >0 *Escherichia coli* (CFU/100ml) - Yes (Ref: No)** | 379 | 302 (80%) | 131 (43%) | 0.13 | -0.38 | 0.64 | 0.62 | 270 (89%) | 1.44 | -1.65 | 3.81 | 0.25 |
| **Drinking water with >10 *TFC* (CFU/100ml) - Yes (Ref: No)** | 379 | 372 (98%) | 159 (43%) | 0.00 | -1.53 | 1.63 | 1.00 | 335 (90%) | 0.41 | -2.54 | 2.22 | 0.71 |
| **Prepared food given to child - Yes (Ref: No)** | 197 | 82 (42%) | 39 (48%) | -0.25 | -0.82 | 0.31 | 0.38 | 77 (94%) | 0.91 | -0.07 | 2.06 | 0.09 |
| **Additions to food before/during cooking (Ref: Additions not added)** |  |  |  |  |  |  |  |  |  |  |  |  |
| Water | 197 | 164 (83%) | 89 (54%) | 0.73 | -0.03 | 1.53 | 0.06 | 141 (91%) | 0.70 | -0.33 | 1.66 | 0.16 |
| Milk | 197 | 30 (15%) | 15 (50%) | -0.06 | -0.84 | 0.72 | 0.88 | 10 (91%) | 0.19 | -1.54 | 3.12 | 0.86 |
| Tea | 197 | 10 (5%) | 5 (50%) | -0.05 | -1.36 | 1.26 | 0.93 | 2 (67%) | -1.47 | -3.86 | 1.62 | 0.24 |
| Nothing | 197 | 28 (14%) | 10 (36%) | -0.74 | -1.61 | 0.07 | 0.08 | 23 (82%) | -0.73 | -1.78 | 0.46 | 0.19 |
| **Food additions heated - Yes (Ref: No)** | 197 | 77 (39%) | 40 (52%) | 0.04 | -0.53 | 0.62 | 0.88 | 69 (90%) | 0.05 | -0.87 | 1.02 | 0.92 |
| **HWWS before preparing food - Yes (Ref: No)** | 197 | 42 (21%) | 24 (57%) | 0.30 | -0.38 | 1.00 | 0.39 | 37 (88%) | -0.16 | -1.17 | 1.01 | 0.77 |
| **Utensils washed with soap and water before preparation - Yes (Ref: No)** | 197 | 30 (15%) | 18 (60%) | 0.42 | -0.36 | 1.23 | 0.30 | 28 (93%) | 0.59 | -0.73 | 2.46 | 0.45 |
| **Surfaces washed with soap and water before preparation - Yes (Ref: No)** | 197 | 27 (14%) | 14 (52%) | 0.03 | -0.79 | 0.85 | 0.95 | 25 (93%) | 0.45 | -0.87 | 2.33 | 0.56 |
| **Preparation surface (Ref: No)** | 197 |  |  |  |  |  |  |  |  |  |  |  |
| Bowl |  | 2 (1%) | 1 (50%) | Ref. |  |  |  | 2 (100%) | Ref. |  |  |  |
| Cooking pot |  | 119 (60%) | 61 (51%) | 0.05 | -3.20 | 3.30 | 0.97 | 107 (90%) | Does not converge | | | |
| Cooking pot (ground) |  | 44 (22%) | 19 (43%) | -0.27 | -3.55 | 3.00 | 0.85 | 41 (93%) | Does not converge | | | |
| Ground |  | 23 (12%) | 15 (65%) | 0.63 | -2.69 | 3.95 | 0.67 | 20 (87%) | Does not converge | | | |
| Stove |  | 4 (2%) | 3 (75%) | 1.10 | -2.66 | 5.14 | 0.55 | 4 (100%) | 0.00 | -94.97 | 94.97 | 1.00 |
| Bed |  | 1 (1%) | 1 (100%) | Does not converge | |  |  | 1 (100%) | 0.00 | -134.31 | 134.31 | 1.00 |
| Table |  | 4 (2%) | 1 (25%) | -1.10 | -5.14 | 2.66 | 0.55 | 1 (25%) | Does not converge | | | |
| **Additions to food after cooking (Ref: Additions not added)** |  |  |  |  |  |  |  |  |  |  |  |  |
| Water | 197 | 88 (45%) | 49 (49%) | -0.23 | -0.79 | 0.33 | 0.43 | 78 (89%) | -0.13 | -1.05 | 0.79 | 0.77 |
| Milk | 197 | 30 (15%) | 26 (54%) | 0.15 | -0.50 | 0.81 | 0.64 | 27 (90%) | 0.08 | -1.08 | 1.58 | 0.90 |
| Tea | 197 | 1 (1%) | 6 (50%) | -0.05 | -1.25 | 1.14 | 0.93 | 1 (100%) | Does not converge | | | |
| Nothing | 197 | 78 (40%) | 41 (52%) | 0.04 | -0.53 | 0.61 | 0.89 | 70 (90%) | 0.07 | -0.85 | 1.04 | 0.88 |
| **HWWS of caregiver before feeding - Yes (Ref: No)** | 197 | 40 (20%) | 21 (52%) | 0.06 | -0.63 | 0.76 | 0.86 | 35 (88%) | -0.23 | -1.24 | 0.94 | 0.67 |
| **HWWS of child before feeding - Yes (Ref: No)** | 197 | 37 (19%) | 19 (51%) | 0.00 | -0.71 | 0.73 | 0.99 | 32 (86%) | -0.34 | -1.36 | 0.83 | 0.53 |
| **Utensils washed before feeding - Yes (Ref: No)** | 197 | 183 (93%) | 97 (53%) | 1.32 | 0.10 | 2.84 | 0.05 | 164 (90%) | 0.45 | -1.46 | 1.86 | 0.58 |
| **Surface washed before feeding - Yes (Ref: No)** | 197 | 123 (62%) | 69 (56%) | 0.52 | -0.06 | 1.10 | 0.08 | 112 (91%) | 0.46 | -0.46 | 1.38 | 0.32 |
| **Who is involved in child feeding** | 197 |  |  |  |  |  |  |  |  |  |  |  |
| Caregiver |  | 150 (76%) | 68 (45%) | Ref. |  |  |  | 133 (89%) | Ref. |  |  |  |
| Another adult |  | 15 (8%) | 6 (40%) | -0.22 | -1.35 | 0.85 | 0.69 | 12 (80%) | -0.67 | -1.94 | 0.88 | 0.33 |
| Child |  | 32 (16%) | 27 (84%) | 1.87 | 0.94 | 3.00 | 0.00 | 31 (97%) | 1.38 | -0.26 | 4.29 | 0.19 |
| **Child seat** | 197 |  |  |  |  |  |  |  |  |  |  |  |
| Bare floor |  | 99 (50%) | 51 (52%) | Ref. |  |  |  | 91 (92%) |  |  |  |  |
| Mat on the floor |  | 62 (31%) | 30 (48%) | -0.13 | -0.76 | 0.51 | 0.70 | 56 (90%) | -0.20 | -1.30 | 0.96 | 0.73 |
| Caregiver’s knee |  | 31 (16%) | 16 (52%) | 0.00 | -0.81 | 0.82 | 0.99 | 24 (77%) | -1.20 | -2.32 | -0.07 | 0.03 |
| Highchair |  | 5 (3%) | 4 (80%) | 1.33 | -0.63 | 4.32 | 0.24 | 5 (100%) | Does not converge | | | |
| **Feeding (Ref: Feeding method not used)** |  |  |  |  |  |  |  |  |  |  |  |  |
| Separate plate/bowl | 197 | 100 (51%) | 46 (46%) | -0.43 | -1.00 | 0.13 | 0.13 | 86 (88%) | -0.33 | -1.27 | 0.58 | 0.47 |
| Cooking vessel | 197 | 7 (4%) | 1 (14%) | -1.90 | -4.85 | -0.10 | 0.08 | 4 (100%) | Does not converge | | | |
| Caregiver hands | 197 | 47 (24%) | 27 (57%) | 0.33 | -0.33 | 1.00 | 0.33 | 38 (93%) | 0.50 | -0.65 | 1.99 | 0.44 |
| Spoon | 197 | 5 (3%) | 2 (40%) | -0.47 | -2.51 | 1.35 | 0.61 | 1 (25%) | -3.37 | -6.41 | -1.26 | <0.01 |
| Child hand | 197 | 71 (36%) | 40 (56%) | 0.32 | -0.26 | 0.91 | 0.29 | 47 (94%) | 0.78 | -0.36 | 2.27 | 0.23 |
| **Liquid given to child (Ref: No)** |  |  |  |  |  |  |  |  |  |  |  |  |
| Nothing | 197 | 18 (9%) | 10 (56%) | 0.19 | -0.79 | 1.19 | 0.70 | 16 (89%) | -0.05 | -1.41 | 1.84 | 0.95 |
| Breastmilk | 197 | 94 (48%) | 48 (51%) | -0.02 | -0.58 | 0.54 | 0.96 | 82 (92%) | 0.56 | -0.37 | 1.57 | 0.25 |
| Water | 197 | 164 (83%) | 88 (54%) | 0.58 | -0.18 | 1.36 | 0.14 | 139 (90%) | 0.41 | -0.68 | 1.38 | 0.43 |
| Animal milk | 197 | 27 (14%) | 17 (63%) | 0.55 | -0.27 | 1.42 | 0.19 | 14 (93%) | 0.55 | -1.14 | 3.47 | 0.61 |
| Tea | 197 | 38 (19%) | 20 (53%) | 0.07 | -0.64 | 0.78 | 0.85 | 23 (88%) | -0.10 | -1.28 | 1.40 | 0.88 |
| Juice | 197 | 20 (10%) | 11 (55%) | 0.17 | -0.76 | 1.12 | 0.73 | 8 (89%) | -0.05 | -1.82 | 2.90 | 0.96 |
| Soup | 197 | 18 (9%) | 5 (28%) | -1.10 | -2.27 | -0.08 | 0.04 | 9 (90%) | 0.07 | -1.67 | 3.02 | 0.94 |
| Powdered milk | 197 | 3 (2%) | 2 (67%) | 0.65 | -1.71 | 3.73 | 0.60 | 1 (50%) | -2.17 | -5.43 | 1.09 | 0.13 |
| **Drinking cup cleaned before feeding - Yes (Ref: No)** | 197 | 189 (96%) | 99 (52%) | 1.19 | -0.30 | 3.13 | 0.15 | 170 (90%) | 1.09 | -0.87 | 2.64 | 0.20 |
| **Raw and cooked food stored separately - Yes (Ref: No)** | 197 | 128 (65%) | 66 (52%) | 0.03 | -0.55 | 0.62 | 0.91 | 114 (89%) | -0.08 | -1.10 | 0.85 | 0.86 |
| **Animal was present in child feeding area during mealtime (Ref: No exposure)** |  |  |  |  |  |  |  |  |  |  |  |  |
| Any animal | 197 | 89 (45%) | 55 (62%) | 0.78 | 0.21 | 1.36 | 0.01 | 83 (93%) | 0.80 | -0.15 | 1.87 | 0.11 |
| Chicken | 197 | 80 (41%) | 49 (61%) | 0.68 | 0.11 | 1.27 | 0.02 | 74 (92%) | 0.60 | -0.36 | 1.67 | 0.24 |
| Cattle | 197 | 10 (5%) | 6 (60%) | 0.37 | -0.91 | 1.76 | 0.57 | 10 (100%) | Does not converge | | | |
| Goat/Sheep | 197 | 30 (15%) | 22 (73%) | 1.12 | 0.29 | 2.04 | 0.01 | 27 (90%) | 0.08 | -1.08 | 1.58 | 0.90 |
| Dog | 197 | 19 (10%) | 11 (58%) | 0.30 | -0.65 | 1.29 | 0.54 | 18 (95%) | 0.82 | -0.84 | 3.74 | 0.43 |
| Cat | 197 | 2 (1%) | 92 (74%) | Does not converge | | | | 2 (100%) | Does not converge | | | |
